# Thyroid cancer polygenic risk score combined with deep learning analysis of ultrasound images improves the classification of thyroid nodules as benign or malignant

**DOI:** 10.1101/2023.04.11.23288041

**Authors:** Nikita Pozdeyev, Manjiri Dighe, Martin Barrio, Christopher Raeburn, Harry Smith, Matthew Fisher, Sameer Chavan, Nicholas Rafaels, Jonathan A. Shortt, Meng Lin, Michael G. Leu, Toshimasa Clark, Carrie Marshall, Bryan R. Haugen, Devika Subramanian, Regeneron Genetics Center, Kristy Crooks, Christopher Gignoux, Trevor Cohen

**Author notes:** Corresponding authors: Nikita Pozdeyev and Christopher Gignoux; Address: 1890 N Revere Ct. MS F600, Aurora, CO 80045.

## Abstract

Evaluating thyroid nodules to rule out malignancy is a very common clinical task. Image-based clinical and machine learning risk stratification schemas rely on the presence of thyroid nodule high-risk sonographic features. However, this approach is less suitable for diagnosing malignant thyroid nodules with a benign appearance on ultrasound. In this study, we developed thyroid cancer polygenic risk scoring (PRS) to complement deep learning analysis of ultrasound images. When the output of the deep learning model was combined with thyroid cancer PRS and genetic ancestry estimates, the area under the receiver operating characteristic curve (AUROC) of the benign vs. malignant thyroid nodule classifier increased from 0.83 to 0.89 (DeLong, p-value = 0.007). The combined deep learning and genetic classifier achieved a clinically relevant sensitivity of 0.95, 95 CI [0.88-0.99], specificity of 0.63 [0.55-0.70], and positive and negative predictive values of 0.47 [0.41-0.58] and 0.97 [0.92-0.99], respectively. An improved AUROC was consistent in ancestry-stratified analysis in Europeans (0.83 and 0.87 for deep-learning and deep learning combined with PRS classifiers, respectively). An elevated PRS was associated with a greater risk of thyroid cancer structural disease recurrence (ordinal logistic regression, p-value = 0.002). This study demonstrates that augmenting ultrasound image analysis with PRS improves diagnostic accuracy, paving the way for developing the next generation of clinical risk stratification algorithms incorporating inherited risk for developing thyroid malignancy.

## Introduction

Thyroid nodules are very common and detected by thyroid ultrasound in up to 65% of the general population, with prevalence increasing in women, with age, and after radiation exposure^1, 2^. Excluding malignancy when evaluating thyroid nodules is important to clinicians and patients. Diagnostic thyroid/neck US is performed in patients with a suspected thyroid nodule, and fine needle aspiration biopsy (FNA) is the procedure of choice to exclude malignancy^3^. It has been estimated that >600,000 FNAs are performed in the United States each year^4^. These FNAs have a high cost to the healthcare system and are associated with mild but relatively frequent clinical complications^5^.

The American Thyroid Association (ATA)^3^ and the American College of Radiology (ACR, TI-RADS)^6^ have developed algorithms that help clinicians to decide which nodules should be biopsied. Both systems rely on the presence of benign or suspicious sonographic features, such as nodule composition, echogenicity, shape, margins, presence of echogenic foci, and nodule size^3, 7, 8^. Due to reliance on suspicious findings on the ultrasound, clinical risk stratification schemas are less useful for thyroid cancer subtypes, such as follicular thyroid cancer (FTC), follicular variant of papillary thyroid cancer (FV-PTC), and Hurthle cell thyroid cancer (HCTC), that may lack classic features of papillary thyroid cancer (PTC) such as hypoechogenicity and microcalcifications^9–11^. To avoid misdiagnosing these cancers, current clinical guidelines^3, 6^ recommend biopsying thyroid nodules with low-risk sonographic appearance greater than 1.5-2.5 cm despite the low probability of cancer in these lesions (5-10%). Consequentially, many biopsies are performed on benign nodules and are potentially avoidable. Only 7-13% of biopsy results fit cytologic categories requiring thyroid surgery (Bethesda categories V and VI); 44-72% produce benign results, and the remaining 9-32% are either inadequate or indeterminate necessitating repeat procedures or expensive molecular testing^12–15^. Therefore, better methods for risk stratification of thyroid nodules are needed.

One path toward reducing unnecessary biopsies involves deep learning methods. Many groups are working on developing computer-assisted diagnosis systems (CAD) employing convolutional neural networks (CNN – a type of deep learning model used in image processing) trained on thyroid ultrasound images to help clinicians with thyroid cancer risk assessments ^16–20^. Such systems perform comparably to expert radiologists for diagnosing malignant thyroid nodules and do not suffer from interobserver variability affecting clinical schemas^21, 22^. However, similar to clinical risk stratification schemas, these CAD systems perform poorly on malignant thyroid nodules lacking suspicious sonographic features (^23^ and this study). This limitation indicates a need to consider other indicators of risk of malignancy independent of these sonographic features.

Population-based registries suggest that thyroid cancer is highly heritable, although the exact genetic components are not well-defined. Genetic effects are estimated to contribute 28-53% to the susceptibility to thyroid cancer^24, 25^. In a study of the Swedish Family-Cancer Database, the tetrachoric correlation for siblings ranged from 0.34–0.51, whereas in non-genetically related spouses, it was 0.04^24^. In the Icelandic Cancer Registry, the relative risk of thyroid cancer in first-degree relatives of an affected individual was 3.02, and it remained significantly elevated up to third-degree relatives. In contrast, no increased risk was observed in unrelated spouses^26^.

*We hypothesize that taking into account an inherited risk of thyroid cancer in combination with ultrasound-based risk assessment will improve thyroid nodule classification.* To test this hypothesis, we trained a deep learning convolutional neural network classifier of thyroid nodules (CNN classifier) using a well-curated database of ultrasound images. We estimated thyroid cancer polygenic risk score (PRS) for ∼ 74K participants in the Colorado Center for Personalized Medicine (CCPM) Biobank^27^. We evaluated the performance of the CNN classifier alone and in combination with PRS, genetic ancestry, and clinical and demographic covariates.

## Materials and Methods

### Thyroid nodule ultrasound images

The electronic health records from the University of Washington (UW, training and validation data set) and the University of Colorado (CU, test data set) healthcare systems were searched for patients that met the following inclusion criteria: 1) Diagnosis of a thyroid nodule or thyroid cancer; 2) High-quality thyroid ultrasound images obtained with a high frequency (10-17 MHz) ultrasonic transducer available in the picture archiving and communication system (PACS). 3) Thyroid nodule diameter of ≥ 10 mm in at least one dimension; 4) A definitive diagnosis for the thyroid nodule established by either histopathology following thyroid surgery or by fine needle aspiration biopsy (FNA) cytology; 5) Genotype information available in the CCPM Biobank (for CU data only) ^27^. Each nodule was assigned an arbitrary identifier in the format VTBxxxx_TNxxxx and the mapping data linking these identifiers to personal identifiable information was only available to the members of the research group involved in the data collection.

Raw ultrasound studies in DICOM format were copied to a secure server and processed with the help of a custom DICOM format parser and image cropping tool. For each nodule, anonymized images in transverse and longitudinal planes and transverse and longitudinal video clips (if available) were collected. Static images and video clips were cropped to remove artifacts, such as embedded text and scale marks, focusing on the region of interest containing the thyroid nodule. Other artifacts, such as caliper marks in a small subset of images, were manually removed using the GIMP software clone tool (GNU Image Manipulation, The GIMP Development team).

### Clinical data

Clinical data was abstracted from the EHR and included nodule dimensions in 3 planes, location within the thyroid (left, right or isthmus), diagnosis established by histopathology or FNA, and radiology, cytology, and surgical pathology reports (Supplementary Tables 1 and 2). TI-RADS points and scores were extracted from radiology reports if documented or assigned as described in the ACR TI-RADS white paper^6^, but only if the report contained sufficient information. Because we were interested in the performance of the TI-RADS system as used by clinicians in routine clinical practice, TI-RADS points and categories were neither reassigned by the authors nor added based on the review of US images. The recommendation to proceed with FNA was estimated based on the TI-RADS features/categories (as extracted from the clinical radiologist reports) and thyroid nodule size^6^, and standard performance metrics were calculated. Ninety-five percent confidence intervals were estimated using a bias-corrected and accelerated (BCa) bootstrap procedure.

**Figure 1.**
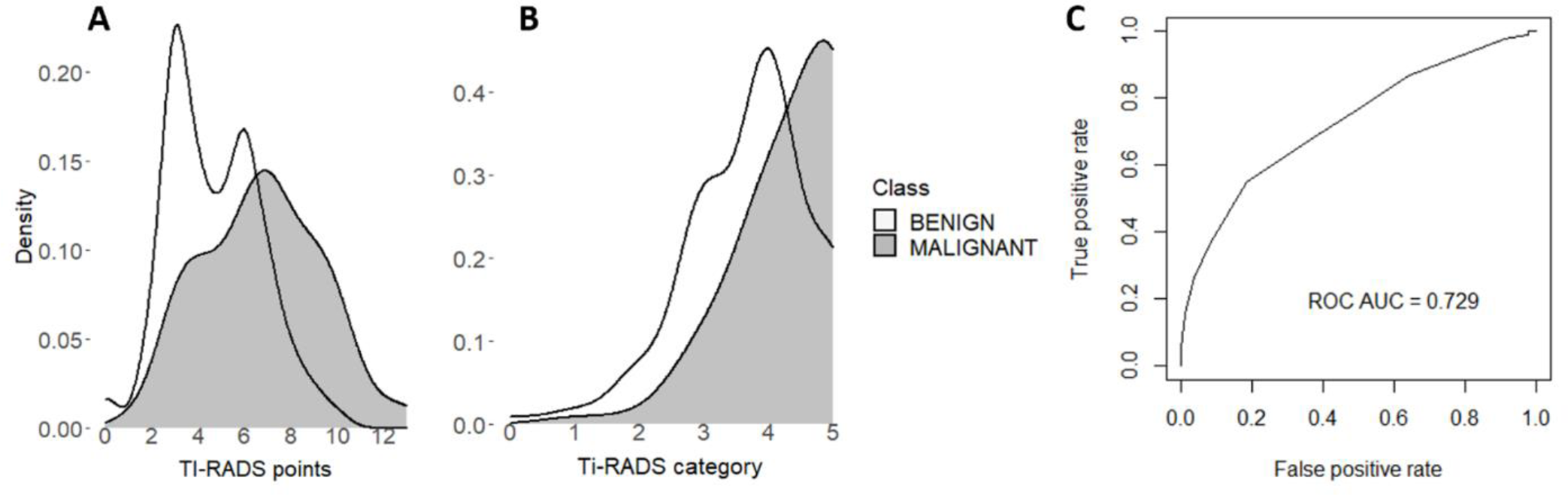
The diagnostic performance of TI-RADS as reported by radiologists in routine clinical practice. Kernel density plots for TI-RADS points (A) and categories (B) and ROC for thyroid nodule classifier based on TI-RADS points (C).

### Thyroid cancer risk of structural disease recurrence

The risk of structural disease recurrence as defined by the ATA thyroid cancer guidelines^3^ (ATA RoR) was estimated for genotyped patients in the CCPM Biobank who underwent total or completion thyroidectomy. A semi-automated pipeline programmed in M using text mining with regular expressions was employed to extract relevant information from the surgical pathology reports. Missing information was curated by a manual chart review by a research team member specializing in thyroid cancer/endocrine surgery (MB). For cases where ATA RoR assignment was ambiguous (such as when the number of nodules with extranodal extension was not reported), the slides were requested from the clinical archive and reviewed by a pathologist with expertise in thyroid cyto- and histopathology (CM). Each malignant specimen was classified as low, intermediate, or high risk for structural disease recurrence (Supplementary Table 3).

### Deep learning thyroid nodule classifier

Thyroid nodule images collected at the UW were used for deep learning thyroid nodule classifier training. The nodules were split into training (80%) and validation (20%) sets, and five-fold cross-validation was used for training and hyperparameter optimization. CNN implementation was done using Python 3.6 and Pytorch 1.9.0. The Big Transfer CNN model^28^ pre-trained on natural images and adopted for thyroid nodule classification through the application of transfer learning was used. The BiT-M ResNet-50×1 CNN architecture was modified to contain 3, 4, 6, and 3 PreActBottleneck units per block 1 through 4. The model trained on the ImageNet-21k dataset^29^ was loaded, and weights for blocks 3 and 4 were fine-tuned for the binary classification task of distinguishing benign and malignant thyroid nodules. A categorical cross-entropy loss function and stochastic gradient descent optimizer were used. The learning rate was increased from zero to 5e-5 during the first 100 epochs (gradual warmup^30^), and training was continued at this learning rate for up to 600 epochs. Training was stopped when the model performance on the validation split did not improve in subsequent epochs. Sample training and validation loss curves are shown in Supplementary Figure 1. The AUROC was calculated for each cross-validation split and summarized using predictions for the held-out component of each split. The optimal scaling, image augmentation, and other hyperparameters were selected using a grid search across the UW training image set. The trained model with optimized parameters was then applied to the CU test set. GradCAM class activation heatmaps^31^ highlighting salient parts of the thyroid nodule image were generated.

Two approaches to image scaling were used: 1) A conventional approach of scaling all images to 400 px regardless of the nodule size. 2) Alternatively, images were scaled based on the reported corresponding dimension of the nodule (100 px per centimeter up to 400 px) with zero padding for nodules <4 cm. Nodules that were >4 cm were scaled to 400 px. Random horizontal flip, scaling (0.8 to 1.2), and shearing (-3 to 3) were applied to all images during each training epoch for data augmentation. Other image augmentation techniques, such as rotation, translation and vertical flip, were tested but did not improve performance on validation set of images. To further augment the data, images extracted from ultrasound cine clips were used for training. Specifically, 2 to 8 randomly selected images from thyroid ultrasound cine clips for each nodule were added to the training image set during each epoch to increase the size and diversity of the training data.

### Genetic data

Patients for the Colorado test set of thyroid nodules were randomly selected among the genotyped participants in the CCPM Biobank (n = 73,346)^27^. Genotyping was performed using Illumina’s InfiniumExpanded Multiethnic Genotyping Array (MEGA^EX^) platform or custom exome sequencing panel in collaboration with the Regeneron Genetics Center. Rigorous quality control, imputation (topmed_r2 reference panel), estimation of kinship-adjusted principal components, genetic ancestry inference, and relationship inference were performed as described previously^27^. One-thousand thirty-five patients surgically treated for benign or malignant nodules were selected from CCPM Biobank participants (Supplementary Table 3).

### Thyroid cancer polygenic risk score

Thyroid cancer PRS was calculated as a weighted sum of 5 or 26 alleles significantly associated with thyroid cancer in a genome-wide association study (GWAS) meta-analysis performed by the Global Biobank Meta-Analysis Initiative (GBMI)^32^. Data from multiethnic thyroid cancer GWAS incorporating evidence for both sexes were used. While CCPM took part in the prior analysis, the summary statistics analyzed here are from GBMI data with CCPM left out to avoid overfitting. The variants and weights used to calculate PRS are summarized in Supplementary Table 4.

### Polygenic risk score phenome-wide association study (PRS PheWAS)

Phecodes for 1860 clinical conditions were derived from ICD-9-CM and ICD-10-CM billing codes for 73,346 CCPM Biobank participants using the *PheWAS* R package^33^. Phenotypes with at least 20 cases were included in the analysis. Phecode exclusions were used in the control definitions as recommended by the original authors. AUROC, p-value, and Nagelkerke r^2^ for crude (unadjusted for covariates) thyroid cancer PRS were calculated for each phenotype (*ROCR* and *fmsb* R packages).

Combined CNN and PRS thyroid nodule classifier.

The probability of malignancy estimated by the CNN classifier was combined with PRS with or without genetic principal components (PCs, accounting for genetic ancestry), nodule dimensions, and demographic covariates (age and sex) using cross-validated (5-fold) logistic regression. Genotyping batch was also used as a covariate for all combined models to control for the origin of the data (array vs. augmented exome). The DeLong test was used to assess the significance of AUROC changes. To be of maximal clinical utility, the thyroid nodule classifier should have high sensitivity and negative predictive power. Therefore, we selected the classifier score thresholds that achieve a sensitivity of >95%, matching or exceeding that of the FNA biopsy^34–37^. The confusion matrix was calculated using these clinically relevant thresholds.

### Human subject research

This retrospective study was approved by the University of Washington Institutional Review Board and the University of Colorado Multiple Institutional Review Board.

Code availability.

The computer code for this manuscript can be found on GitHub https://github.com/npozdey/thyroid_nodule_PRS.

## Results

### Database of ultrasound images and video clips of thyroid nodules

Ultrasound images for CNN classifier training were collected for 621 nodules (458 benign and 163 malignant) from the University of Washington healthcare system (Supplementary Table 1). Five hundred fifty-nine nodules had cine clips obtained in a transverse plane, and 213 had cine clips obtained in a sagittal plane (a total of 32,545 images). Four hundred and sixty-six and 155 nodules were from females and males, respectively. The median age of patients at the time of diagnosis was 51 years. Thirty-seven nodules were in the thyroid isthmus, 326 in the right thyroid lobe, and 258 in the left thyroid lobe. Thyroid nodules ranged in size from 1 to 9.4 cm (median = 2.7 cm). Significantly more malignant nodules (41/163, 25%) than benign nodules (76/458, 17%) were taller than wide (Pearson Chi-squared test, p = 0.02).

To approximate better the variety of nodules that clinicians encounter in clinical practice, all thyroid nodule types, including those that originate from non-thyroidal tissues, were used. Two benign nodules were diagnosed as intra-thyroidal parathyroid adenomas. One hundred and twelve thyroid nodules were from PTCs of classic or unspecified subtype. The database also included 19 FV-PTC, 15 FTC, three medullary thyroid carcinomas (MTC), three HCTC, two metastases from neuroendocrine tumors, two metastases from chronic lymphocytic leukemia, two metastases from breast carcinomas, one metastasis from colorectal carcinoma, one diffuse sclerosing variant of PTC, one poorly differentiated thyroid carcinoma, one parathyroid carcinoma, and one thyroid lymphoma.

Test imaging data was acquired from the medical records of patients participating in the CCPM Biobank (Supplementary Table 2). Among 232 nodules, there were 168 benign nodules, 44 classic PTCs, 12 FV-PTC, 5 MTC, 2 FTC, and 1 HCTC.

### TI-RADS has high sensitivity but low specificity when used by radiologists in routine clinical practice

To create a context for assessing CNN and PRS-based thyroid nodule risk stratification, we evaluated TI-RADS schema in our training set of nodules. The radiologists at the UW use TI-RADS consistently, therefore, we were able to extract TI-RADS points and categories from clinical reports for most thyroid nodules (Supplementary Table 1). We argue that this method provides a better estimate of the real-life clinical performance of TI-RADS in contrast to a dedicated research study, where ultrasounds may be reanalyzed by a physician experts, and evaluations by multiple physicians are reconciled (e.g.^38, 39^). Kernel density plots illustrate the ability of TI-RADS points and categories to discriminate between benign and malignant thyroid nodules (Figure 1). The nodules with ≥ 9 TI-RADS points are very likely to be malignant (Figure 1A). However, there is a significant overlap between benign and malignant nodules, particularly for nodules with ≤ 7 points or category of ≤ 4. The AUROCs of TI-RADS points and TI-RADS categories were 0.729 (Figure 1C) and 0.707, respectively. Thyroid nodule size was considered to provide the recommendation to proceed with FNA or use active surveillance as recommended by the TI-RADS white paper^6^. This binary TI-RADS classifier had a sensitivity of 0.93, 95% CI [0.86, 0.97], a specificity of 0.19 [0.15, 0.23], and a negative predictive value (NPV) of 0.91 [0.83, 0.96] and a positive predictive value of 0.23 [0.19, 0.28].

### Deep learning model training and performance

The AUROCs averaged across five cross-validation splits using various image scaling and augmentation techniques are listed in Table 1. When traditional scaling to 400 px (not taking nodule size into account) was used, the model achieved an AUROC of 0.803 (summarized across five independent cross-validated training runs). Scaling by nodule size significantly improved the AUROC to 0.848 (p=0.0001, DeLong test). Additional improvement was achieved when six random frames were extracted from cine clips and added to the training set during each epoch (AUROC 0.872, p = 0.0009 when compared to the model trained on images scaled by the nodule size). Supplementary Figure 2 shows the ROC curve (AUROC 0.86) from the example training run using both scaling of the images by the nodule size and extracting random 6 video clip frames per nodule during each epoch. When the probability of malignancy threshold (*P*_malign_ *≥ 0.07)* was set to match the sensitivity of the FNA, the CNN thyroid nodule classifier showed a sensitivity of 0.95 [95% CI, 0.91-0.98], a specificity of 0.52 [0.48-0.57], an NPV of 0.97 [0.94-0.98], and a positive predictive value (PPV) of 0.41 [0.36-0.46] (Table 2). When evaluated on the test set of 232 nodules, the AUROC was 0.833 (Table 2).

**Table 1.** Cross-validated AUROC of CNN thyroid nodule classifier trained using various image scaling and augmentation methods.

| Image scaling | Image augmentation | AUROC | P-value |
| --- | --- | --- | --- |
| Scaled to 400 px | Standard* | 0.804 ± 0.007 | |
| By nodule size | Standard | 0.848 ± 0.005 | 0.0001<sup>§</sup> |
| By nodule size | Six random cine clip images added | 0.872 ± 0.004 | 0.0009<sup>§§</sup> |

\*Standard image augmentation consists of random horizontal flip ( $p = 0.5$ ), random scaling (0.8 to 1.2) and random shear $(-3, 3)$ . <sup>s</sup>When compared to scaling to 400 px, DeLong test. <sup>ss</sup>When compared to scaling by nodule size. Data is shown as mean ± standard deviation. N = 5.

**Table 2.** AUROCs, diagnostic, and predictive metrics of thyroid nodule classifiers.

|  | AUROC,<br>p-value<sup>‡</sup> | Sensitivity*<br>[95% CI] | Specificity<br>[95% CI] | PPV<br>[95% CI] | NPV<br>[95% CI] |
| --- | --- | --- | --- | --- | --- |
| CNN, training data<sup>†</sup> | 0.87 | 0.95<br>[0.91 -0.98] | 0.52<br>[0.48-0.57] | 0.41<br>[0.36-0.46] | 0.97<br>[0.94-0.98] |
| CNN, test data | 0.83 | 0.89<br>[0.79-0.95] | 0.62<br>[0.54-0.69] | 0.47<br>[0.38-0.56] | 0.94<br>[0.88-0.97] |
| CNN + PRS<sup>†</sup> | 0.87, 0.05 | 0.95<br>[0.88-0.99] | 0.57<br>[0.49-0.64] | 0.46<br>[0.38-0.54] | 0.97<br>[0.92-0.99] |
| CNN + PRS + PCs<sup>†</sup> | 0.89, 0.007 | 0.95<br>[0.88-0.99] | 0.63<br>[0.55-0.70] | 0.47<br>[0.41-0.58] | 0.97<br>[0.92-0.99] |
| CNN + PRS +<br>covariates<sup>†</sup> | 0.92, 2.3e-4 | 0.95<br>[0.88-0.99] | 0.56<br>[0.48-0.63] | 0.45<br>[0.37-0.53] | 0.97<br>[0.91-0.99] |
| CNN, test data,<br>Europeans only | 0.83 | 0.87<br>[0.75-0.94] | 0.62<br>[0.54-0.70] | 0.47<br>[0.38-0.56] | 0.92<br>[0.84-0.97] |
| CNN + PRS<sup>†</sup>,<br>Europeans only | 0.87, 0.03 | 0.96<br>[0.87-1.00] | 0.58<br>[0.50-0.66] | 0.50<br>[0.40-0.59] | 0.97<br>[0.90-1.00] |

\*Binary classifier threshold was selected to achieve clinically relevant sensitivity of ~ 0.95; <sup>†</sup>5-
fold cross-validated; <sup>‡</sup>compared to CNN model evaluated on the test set of nodules; CNN –
convolutional neural network classifier; PRS – 5 SNP PRS; PC – genetic principal component

### Malignant and benign image characteristics recognized by the CNN classifier

To understand benign and malignant features recognized by the CNN classifier, we explored GradCAM class activation heatmaps highlighting salient areas of the image. Of particular interest were heatmaps of thyroid nodules classified as malignant or benign with high confidence (*P*_malign_ *∼ 1 or 0*, respectively). GradCAM heatmaps for the 16 nodules with the most extreme *P*_malign_ are shown in Fig. 2). Microcalcifications were present in all eight top malignant nodules (#1-8) and were highlighted as a salient feature by GradCAM (Figure 2A). Benign features were the presence of multiple small cystic areas (#9, 10, 13, 15, 16), isoechoic nodules with well-defined borders (#11 and 12), and purely cystic nodule (#14, heatmap points to the cyst borders). Benign nodules #9, 10, 12, 13, and 15 were recommended for FNA based on the clinical ultrasound report and TI-RADS algorithm. Similar benign and malignant features were highlighted on GradCAM heatmaps generated from the test set images (Supplementary Fig. 3). Nodules with the highest *P*_malign_ have classic high-risk appearance due to hypoechogenicity and the presence of microcalcifications (VTB5276_TN5276, VTB5093_TN5093, VTB5327_TN5327, Supplementary Figure 3). Nodules with the low *P*_malign_ demonstrate spongiform architecture (VTB5378_TN5379, VTB5151_TN5151, VTB5375_TN5375) or are solid and isoechoic (VTB5110_TN5110, VTB5291_TN5291, VTB5111_TN5111).

**Figure 2.**
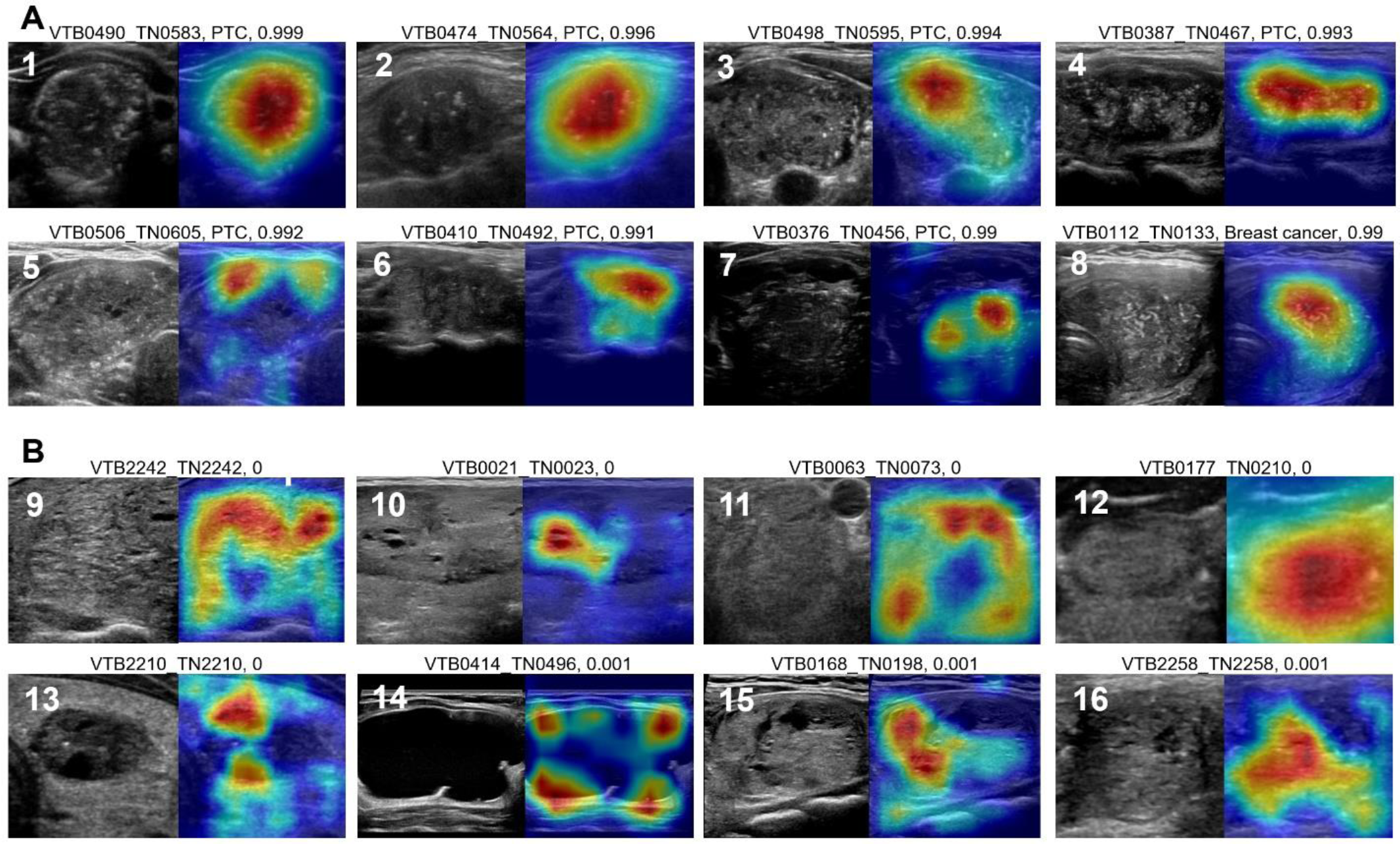
Malignant (A) and benign (B) thyroid nodules classified by CNN model with high cofidence. Nodule ID, the probability of malignancy estimated by the deep learning classifier, US image, and GradCAM activation heatmap for the corresponding class are shown.

### Malignant thyroid nodules misclassified as benign

To understand the limitations of thyroid nodule risk stratification, we explored malignant thyroid nodules assigned low *P*_malign_ of <0.1 (Figure 3A). Notably, none of these nodules demonstrate overtly suspicious sonographic features. Many of the misclassified nodules belong to difficult-to-diagnose subtypes such as FTC (#17), FV-PTC (#21, 22, 26), and intrathyroidal hematologic malignancies (#20 and 24). PTC #19 had benign FNA cytology, yet surgical histopathology showed multifocal microcarcinoma (0.5 and 0.7 cm), suggesting that the image may not represent the malignant tumor. Nodule #25 was described as “oncocytic variant of PTC“ in the histopathology report and may resemble HCTC. Among five misclassified nodules in a test set (Figure 3B), four showed no overt suspicious characteristics (except for HCTC # 30, which is hypoechoic and taller than wider). All five belonged to difficult-to-diagnose types of thyroid cancer: FV-PTC, FTC, and HCTC. This analysis highlights the inherent weakness of image-based thyroid nodule risk stratification (both clinical and machine learning), i.e. the benign sonographic appearance of a subset of malignant lesions. This observation prompted us to seek an orthogonal method of risk assessment, such as leveraging the inherited risk of thyroid cancer by using PRS.

**Figure 3.**
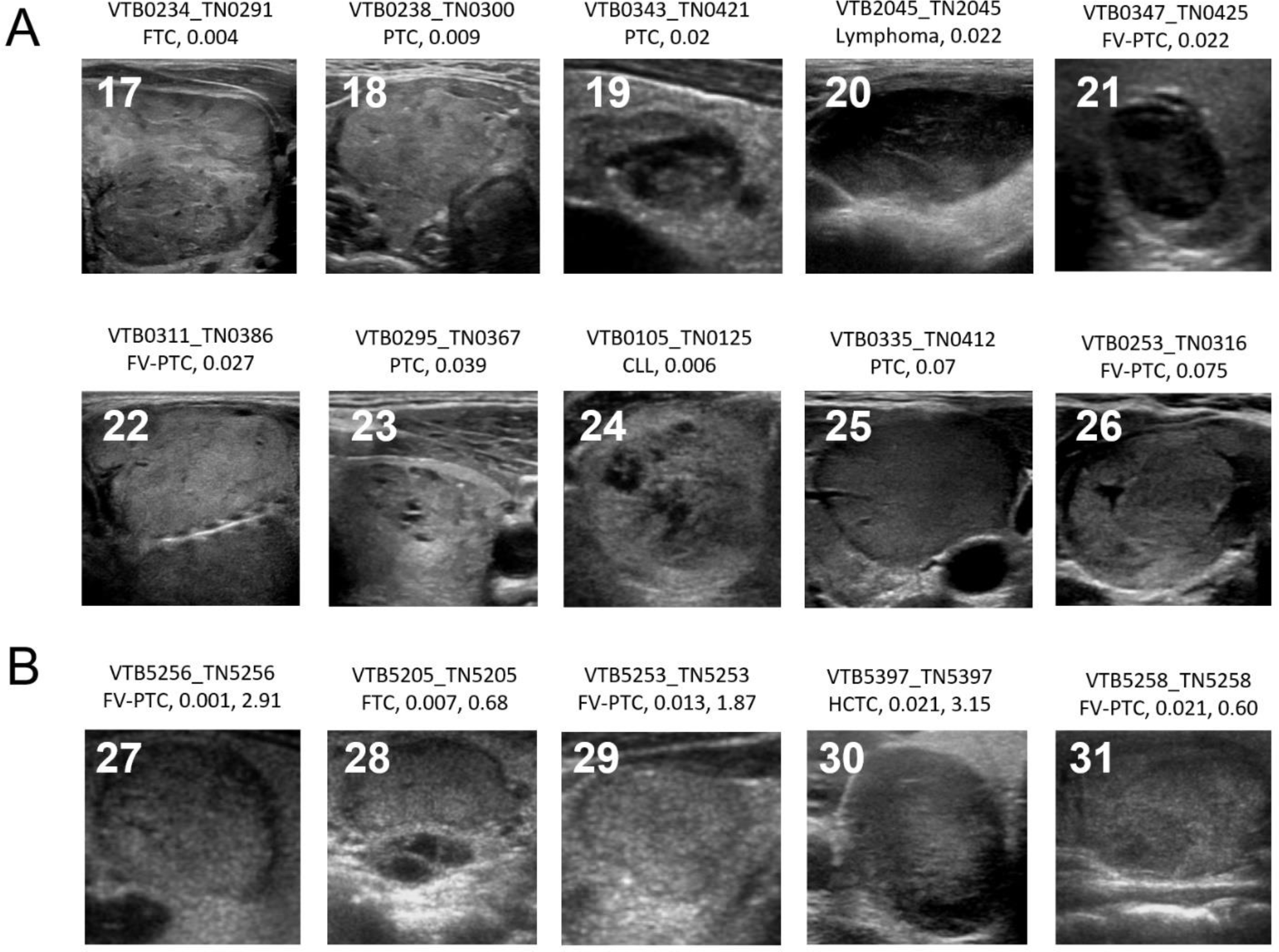
Malignant thyroid nodules incorrectly classified as benign by the deep learning model in a training (A) and test (B) sets of images. Nodule ID, histologic tumor type, and P_malign_ are shown. For test nodules (B), standardized PRS is also shown. CLL – chronic lymphocytic leukemia.

### Thyroid cancer polygenic risk score

Initially, we derived a PRS from 26 variants significantly associated with thyroid cancer in a GBMI meta-analysis (26 SNP PRS) for the Colorado Center for Personalized Medicine Biobank participants. This score achieved an area under the receiver operating characteristic curve of 0.65 in ∼ 74K CCPM Biobank participants (Supplementary Figure 4A) and 0.70 when five genetic principal components, age, sex, and genotyping batch were included as covariates). This AUROC is comparable to that of previously published thyroid cancer PRS^40, 41^. To understand the predictive landscape of the 26 SNP PRS we performed PRS PheWAS. In addition to thyroid cancer, 26 SNP PRS was significantly associated with other thyroid disease phenotypes (such as hypothyroidism, chronic lymphocytic thyroiditis, and nontoxic nodular goiter, Supplementary Figure 4C, Supplementary Table 5). When restricted to individuals of European ancestry, the PRS showed an AUROC of 0.65, similar to the AUROC calculated in the entire CCPM Biobank population (Supplementary Fig. 4B).

The association of 26 SNP PRS with benign nodular goiter is detrimental to its ability to discriminate between malignant and benign nodules (a clinical use case of this study). Therefore, we studied the association of each variant with the phenome using PheWAS (Supplementary Table 6) and found that some variants (i.e. chr1:218508629:G:A and chr8:132869226:C:T) are associated with nontoxic nodular goiter more than with thyroid cancer. To remove the negative effect of the variants associated with the benign nodular goiter on the ability of PRS to discriminate between malignant and benign thyroid nodules (and to avoid overfitting by selecting just the variants with the best performance in the CCPM Biobank), we limited PRS to the five most significant SNPs as discovered by the GBMI (5 SNP PRS, Supplementary Table 4). These variants replicated previous discoveries in much smaller GWASes ^42–46^ and, therefore, are less likely to be affected by an ascertainment bias.

The 5 SNP PRS achieved an AUROC of 0.63 (0.68 when covariates were included) (Figure 4A). PRS PheWAS analysis demonstrated that the 5 SNP PRS retained a strong association with thyroid cancer phenotype but not with nontoxic nodular goiter phenotype (Figure 4B, Supplementary Table 7) making it suitable for the task of thyroid nodule classification.

**Figure 4.**
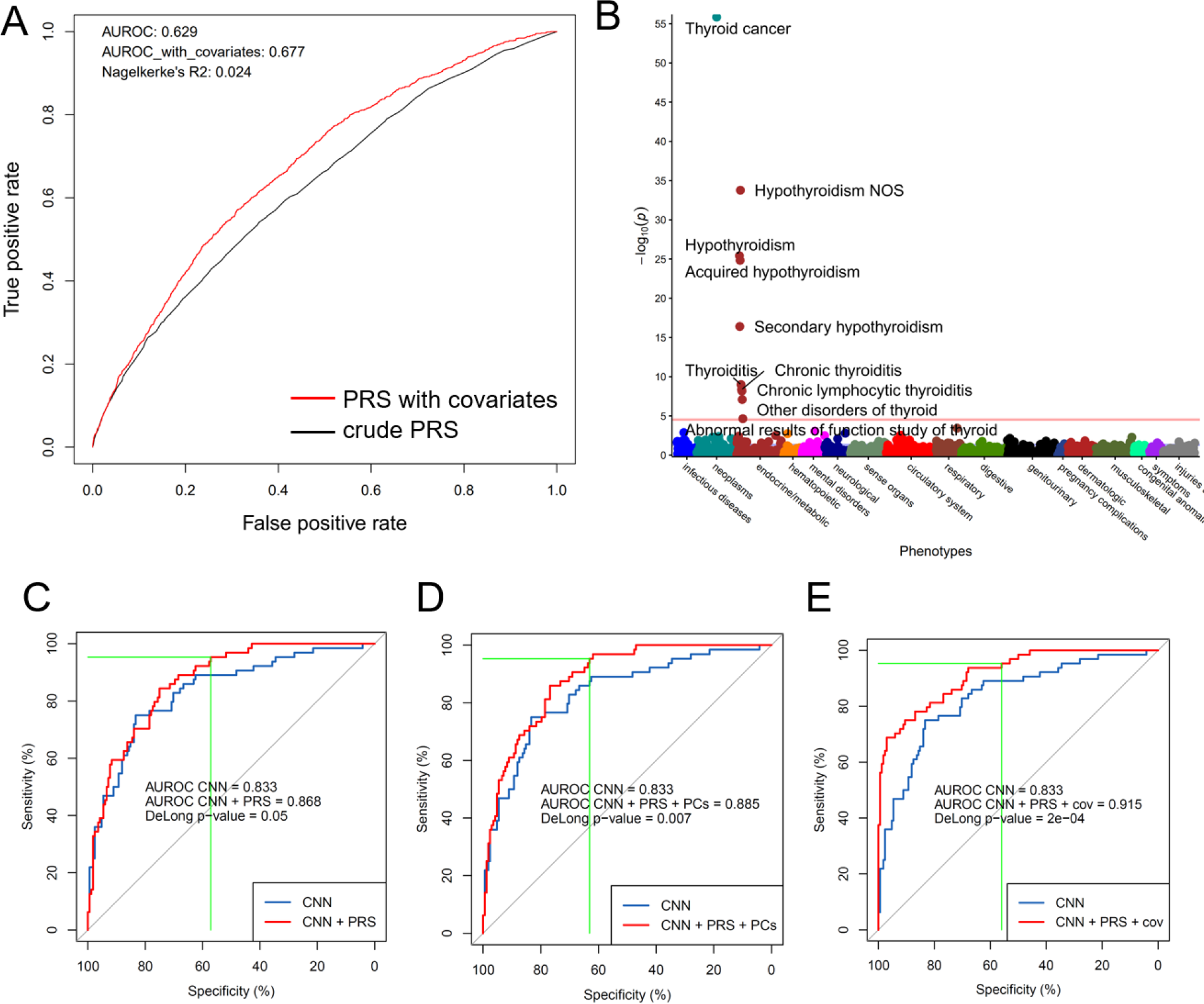
Thyroid cancer 5 SNP PRS alone and in combination with CNN thyroid nodule classifier. A. ROC curve of the 5 SNP PRS in CCPM Biobank participants (blue curve – crude PRS; red curve – PRS with covariates: 5 principal components, genotyping batch, age and sex). B. 5 SNP PRS PheWAS. C. CNN thyroid nodule classifier combined with 5 SNP PRS. D. CNN thyroid nodule classifier combined with 5 SNP PRS and 5 principal components. E. CNN thyroid nodule classifier combined with 5 SNP PRS, 5 principal components, nodule dimensions and age and sex.

### PRS improves the classification of thyroid nodules as benign and malignant and is associated with thyroid cancer risk of recurrence

Combining the CNN output with 5 SNP PRS using a cross-validated logistic regression model increased AUROC from 0.83 to 0.87 (DeLong test, p = 0.05, Figure 4C, Table 2). Adding genetic PCs to the model resulted in further improvement of AUROC to 0.89 (DeLong test, p = 0.007, Fig. 4D, Table 2). Finally, including patient demographics (age, sex) and nodule dimensions as covariates resulted in an AUROC of 0.92 (DeLong test, p = 2-e04, Figure 4E, Table 2).

To investigate the possibility that improved performance of the combined classifier is driven by subtle differences in PRS among individuals of different ancestries (i.e. PRS is a surrogate measure of ancestry and not a true estimate of thyroid cancer risk), we performed a subanalysis including patients of European genetic ancestry only. Similarly to the multiethnic analysis, PRS increased the AUROC of the thyroid nodule classifier from 0.83 to 0.87 (Table 2, De-Long test p = 0.03) in 181 European patients, indicating that this improved performance is not caused by the confounding effect of population structure.

ATA RoR was determined for 842 CCPM Biobank participants operated for thyroid cancer. Clinical charts and slides from surgical histopathology were reviewed as necessary to assign ATA RoR per guidelines^3^ (611 low-risk, 152 intermediate-risk, and 79 high-risk ATA RoR, Supplementary Table 3). Ordinal logistic regression showed a significant positive association of 5 SNP PRS with ATA RoR (β_PRS_ = 0.22, 95% CI [0.08-0.36], p-value = 0.002). In addition, male sex (β_Male_ = 1.01 [0.67-1.35], p-value = 5.68e-9) and self-reported Hispanic ethnicity (β_Hispanic_ = 0.76 [0.24-1.26], p-value = 0.003) were associated with greater risk of structural disease recurrence.

## Discussion

In this study, we investigated the utility of thyroid cancer PRS for two use cases of clinical relevance: thyroid nodule risk stratification necessary for thyroid cancer diagnosis and estimating the risk of thyroid cancer structural disease recurrence, which determines the type of surgical treatment, need for radioactive iodine treatment and the degree of TSH suppression in patients with thyroid malignancy^3^.

A thyroid cancer diagnosis is established with an FNA biopsy of nodules. As it is not feasible nor cost-effective to biopsy all thyroid nodules, the ATA and the ACR introduced clinical risk stratification algorithms^3, 6^ that help to identify nodules that are more likely to be malignant based on the presence of suspicious sonographic characteristics such as hypoechogenicity, presence of microcalcifications, and others. However, not all malignant thyroid nodules demonstrate these suspicious features on ultrasound. In particular, FTC ^11^, FV-PTC ^47^, and HCTC^48^ frequently have benign sonographic appearances limiting the usefulness of image-based risk stratification schemas. To avoid missing benign-appearing thyroid malignancies, ACR and ATA recommend biopsying benign-looking thyroid nodules ≥ 1.5-2.5 cm in size, which leads to many potentially avoidable FNA of benign lesions. Consistently, our analysis of TI-RADS, as it was used by radiologists in routine clinical practice, showed high sensitivity but low specificity for detecting malignancy (Figure 1).

To objectively assess the risk of malignancy based on the analysis of thyroid nodule ultrasound images, we trained a CNN classifier using a diverse and well-annotated set of images. We found that preserving the size of sonographic features using image scaling based on the nodule size and increasing the diversity of training data by pulling a random set of cine clip images during each training epoch had a positive influence on model performance (Table 1). These techniques could be useful for a broad range of medical image analysis applications.

The CNN classifier performed well in distinguishing benign and malignant nodules (AUROC 0.83-0.87), outperforming the TI-RADS risk stratification schema as it was used by clinicians in routine clinical practice (Figure 1, AUROC 0.71-0.73). The CNN classifier recognized many features known to be associated with malignancy (hypoechogenicity and microcalcifications) and benign nodules (spongiform appearance and isoechoic appearance, Figure 2). However, it misclassified nodules that do not have suspicious features on the ultrasound and belong to the hard-to-diagnose subtypes of thyroid cancer such as FTC, HCTC, and some FV-PTC.

Inherited predisposition to developing thyroid cancer measured with PRS provides an attractive alternative method of measuring risk, independent and complementary to the imaged-based assessment. Published data on the clinical utility of thyroid cancer PRS are scarce. Integration of the PRS with clinical factors was shown to improve the prediction of subsequent thyroid cancer in childhood cancer survivors^49^. Individual variants were tested for their association with tumor size, locoregional and distant metastases, extrathyroidal extension, and multifocality (reviewed in ^50^). We demonstrated that incorporating PRS into the thyroid nodule risk stratification results in an improved ability to discriminate between benign and malignant nodules as measured by the classifier AUROC (Table 2). In clinical practice, this may help to reduce the number of potentially avoidable FNAs performed for benign lesions.

Many GWAS analyses identified germline variants associated with thyroid cancer^42–46^, culminating with the largest to-date meta-analysis performed by the GBMI^32^. Our PheWAS and PRS PheWAS analyses discovered that not all variants significantly associated with thyroid cancer in GWAS are suitable for differentiating benign and malignant thyroid nodules due to pleiotropic interaction with benign and malignant phenotypes. There are two potential sources of genetic associations with benign goiter in thyroid cancer GWAS. One is the true shared genetic basis for thyroid cancer and benign nodules. However, thyroid cancer and benign goiter are separate nosological entities, and benign thyroid nodules are not considered premalignant^51^. The other is ascertainment bias due to the greater prevalence of benign thyroid nodules in patients with thyroid cancer^52^ and, importantly, due to the exclusion of benign thyroid disease from the control group but not from the case group in GWAS (such as when using recommended phecode exclusions^33^). To reduce the effect of the ascertainment bias, we limited PRS to the five most significant SNPs and confirmed the association of this sparse PRS with thyroid cancer but not benign nodular goiter in PRS PheWAS (Figure 4). Further discovery of the variants specific to thyroid cancer but not benign nodular disease is needed, which can be achieved through the careful selection of case and control phenotypes in GWAS (such as incorporating patients with benign nodules into the control definition for thyroid cancer GWAS).

The ATA RoR is essential for many aspects of thyroid cancer clinical management, including the need for completion thyroidectomy and adjuvant radioactive iodine treatment. However, ATA RoR, as defined in the current guidelines^3^, relies on surgical histopathology and, therefore, is not known until after the thyroid surgery creating uncertainly in the optimal selection of the initial surgical treatment (lobectomy for low-risk and total thyroidectomy with and without central neck dissection for high-risk disease). We found a significant positive association between PRS and ATA RoR, which is of potential clinical relevance. In addition to PRS, we confirmed an association of male sex (reviewed in ^53^) with higher risk thyroid cancer. Finally, a positive association between Hispanic ethnicity and ATA RoR highlights ethnic disparity in thyroid cancer, which may have genetic or socioeconomic origin and warrants future study.

This study has limitations. The sparse 5 SNP PRS explains a small fraction of heritable thyroid cancer risk, a consequence of our finite understanding of thyroid cancer genetic architecture. We expect that, as our knowledge of genetic variants increasing thyroid cancer risk grows, the performance and clinical utility of PRS will improve. Despite the participation of multiple Biobanks from across the globe in the GBMI^32^, the thyroid cancer meta-analysis was enriched in genetic data from participants of European ancestry. The ancestry-stratified analysis in Europeans confirmed a positive contribution of PRS to thyroid nodule risk assessment (Table 2). However, the utility of this approach in non-European individuals could not be tested given sample size limitations and may be suboptimal^54^. Performing ancestry-specific thyroid GWAS, developing ancestry-adjusted PRS, and performing testing on patients of various races and ethnicities are needed to ensure equitable performance.

In summary, we demonstrated the utility of PRS for the diagnosis of thyroid cancer and the association of PRS with the risk of thyroid cancer recurrence, paving the path for the clinical use of thyroid cancer inherited risk assessment in combination with image analysis. The proposed CNN + PRS thyroid nodule classifier is deterministic and, therefore, does not suffer from interobserver variability. It could be particularly impactful for clinical care in community/rural settings with less expertise in interpreting thyroid ultrasounds and detecting suspicious features of thyroid nodules.

## Grant support

This work was supported by the grant from the University of Colorado Cancer Center to NP and Colorado Cancer League fellowship AWD#222494-MB to MB. Genotyping data was provided by the Colorado Center for Personalized Medicine.

## Disclosures

Bryan R. Haugen received research support from Eisai and Merck unrelated to this research study. Bryan R. Haugen served on an Advisory Board at Eisai and is currently a member of the finance committee for the International Thyroid Oncology Group and Endocrine Society. Regeneron Genetics Center is a subsidiary of Regeneron Pharmaceuticals. All other authors have completed the ICMJE uniform disclosure form and declare: no support from any organization for the submitted work. Regeneron Genetics Center contributed to this study by genotyping the Colorado Center for Personalized Medicine participants and was not involved in the design, data analysis, or manuscript writing.

## Data Availability

All data produced in the present study are either contained in the manuscript or available upon reasonable request to the authors, with the exception of the individual-level genetic data.

https://github.com/npozdey/thyroid_nodule_PRS

## Notes

### Competing Interest Statement

Martin Barrio was supported by the Cancer League of Colorado fellowship WD#222494-MB. Nikita Pozdeyev was supported by a research grant from the University of Colorado Cancer. Bryan R. Haugen receives research support from Eisai and Merck unrelated to this research study. Bryan Haugen served on an Advisory Board at Eisai and is currently a member of the finance committee for the International Thyroid Oncology Group and Endocrine Society. Regeneron Genetics Center is a subsidiary of Regeneron Pharmaceuticals. All other authors have completed the ICMJE uniform disclosure form and declare: no support from any organization for the submitted work.

### Funding Statement

This work was funded by the grant from the University of Colorado Cancer Center to NP and Colorado Cancer League fellowship AWD#222494-MB to MB. Genotyping data was provided by the Colorado Center for Personalized Medicine.

### Author Declarations

IRB of the University of Washington gave ethical approval for this work. Colorado Multiple Institutional Review Board gave ethical approval for this work.

