## Supplementary Figures 1,2,4 for "Thyroid cancer polygenic risk score combined with deep learning analysis of ultrasound images improves the classification of thyroid nodules as benign or malignant"

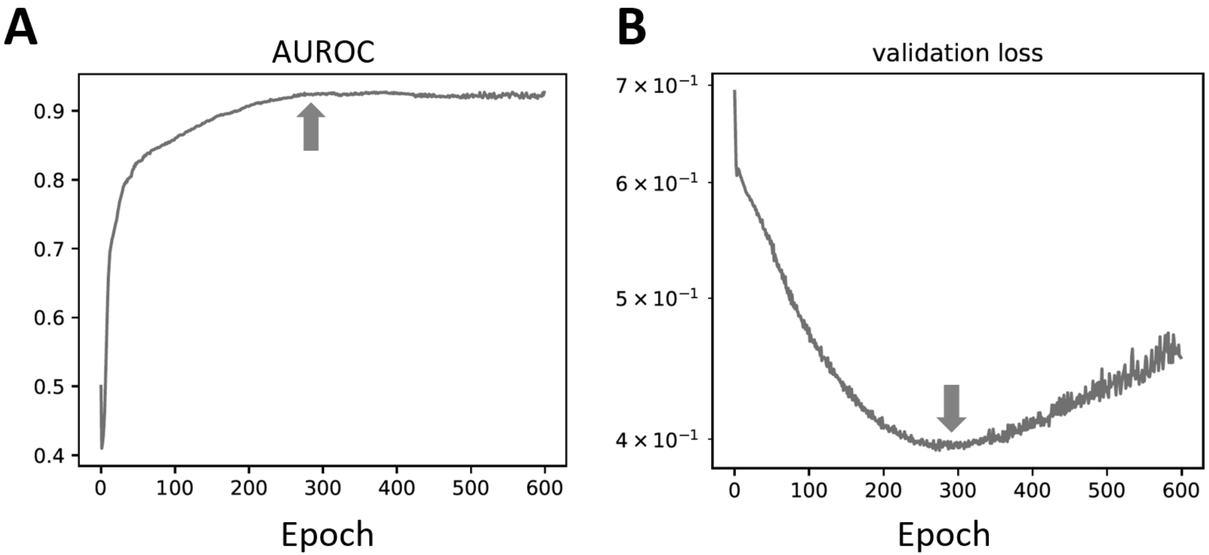


**Supplementary Figure 1. Example training curves for DL thyroid nodule classifier.** Changes in AUROC (A) and validation image set loss (B) with training epochs are shown. In this example, training was continued for 600 epochs, and the model was overfitted on validation images after epoch 290. The arrows show a training epoch when the validation loss reached its minimum.

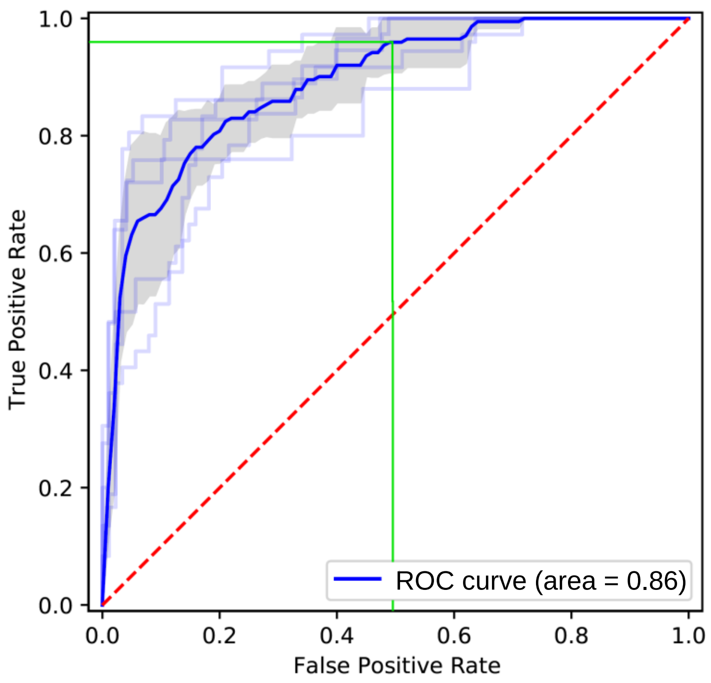


**Supplementary Figure 2. Receiver-operating characteristic curve for deep learning thyroid nodule classifier scaling by nodule size and 4 images per video clip per nodule.** ROC for 5 cross-validation folds and average are shown. Threshold for binary classification was set to match the sensitivity of thyroid nodule FNA biopsy (≥ 0.95).

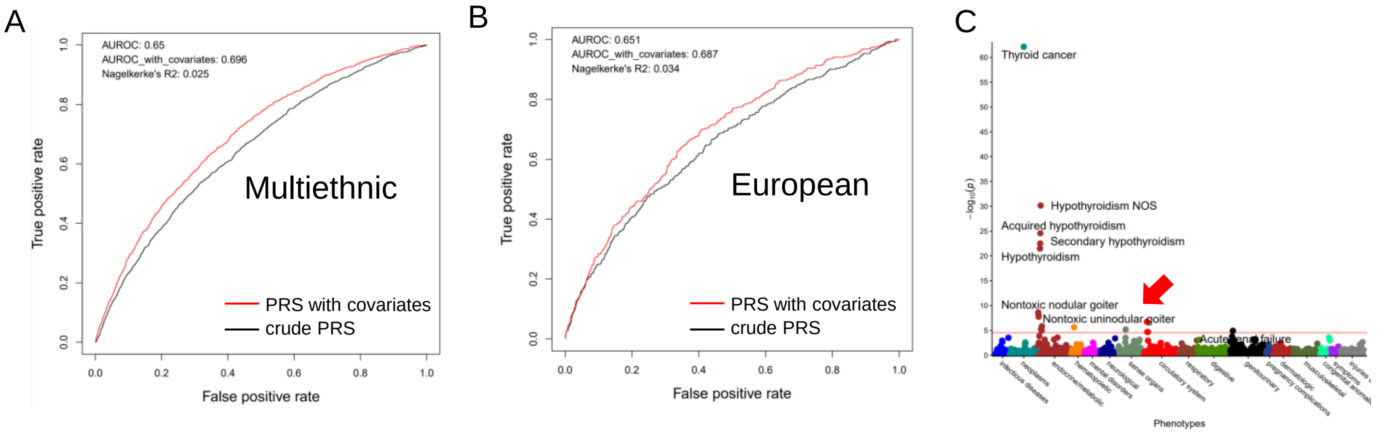


**Supplementary Figure 4. Thyroid cancer PRS ROC curve for the entire CCPM Biobank (A) and a subset of individuals of European ancestry (B). C. 26 SNP PRS PheWAS (red arrow highlights the association of 26 SNP PRS with benign nodular goiter phenotypes).**
