## Supplementary Figure 3 for "Thyroid cancer polygenic risk score combined with deep learning analysis of ultrasound images improves the classification of thyroid nodules as benign or malignant"

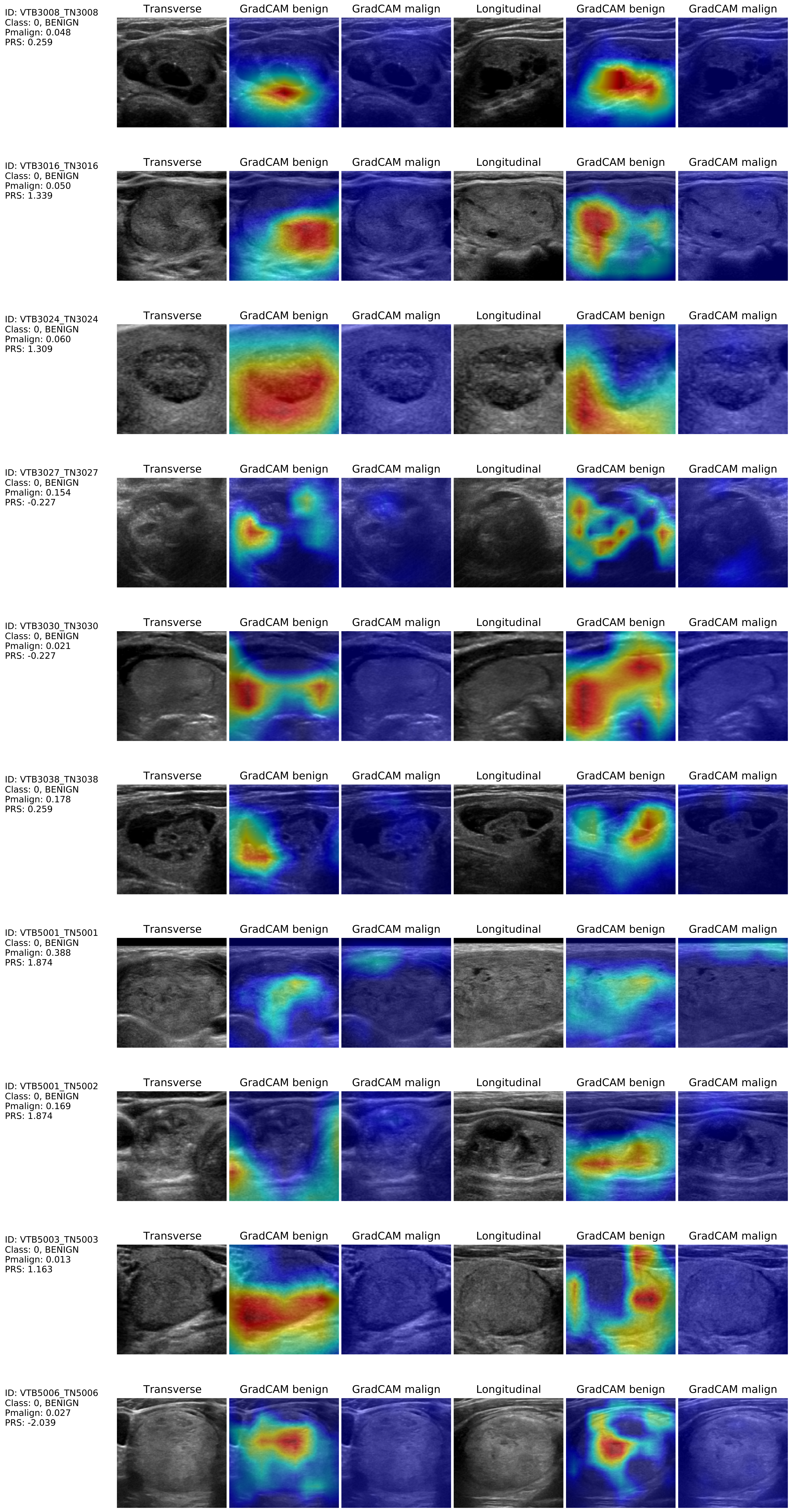
ID: VTB3008_TN3008
Class: 0, BENIGN
Pmalign: 0.048
PRS: 0.259
Transverse
GradCAM benign
GradCAM malign
Longitudinal
GradCAM benign
GradCAM malign
ID: VTB3016_TN3016
Class: 0, BENIGN
Pmalign: 0.050
PRS: 1.339
Transverse
GradCAM benign
GradCAM malign
Longitudinal
GradCAM benign
GradCAM malign
ID: VTB3024_TN3024
Class: 0, BENIGN
Pmalign: 0.060
PRS: 1.309
Transverse
GradCAM benign
GradCAM malign
Longitudinal
GradCAM benign
GradCAM malign
ID: VTB3027_TN3027
Class: 0, BENIGN
Pmalign: 0.154
PRS: -0.227
Transverse
GradCAM benign
GradCAM malign
Longitudinal
GradCAM benign
GradCAM malign
ID: VTB3030_TN3030
Class: 0, BENIGN
Pmalign: 0.021
PRS: -0.227
Transverse
GradCAM benign
GradCAM malign
Longitudinal
GradCAM benign
GradCAM malign
ID: VTB3038_TN3038
Class: 0, BENIGN
Pmalign: 0.178
PRS: 0.259
Transverse
GradCAM benign
GradCAM malign
Longitudinal
GradCAM benign
GradCAM malign
ID: VTB5001_TN5001
Class: 0, BENIGN
Pmalign: 0.388
PRS: 1.874
Transverse
GradCAM benign
GradCAM malign
Longitudinal
GradCAM benign
GradCAM malign
ID: VTB5001_TN5002
Class: 0, BENIGN
Pmalign: 0.169
PRS: 1.874
Transverse
GradCAM benign
GradCAM malign
Longitudinal
GradCAM benign
GradCAM malign
ID: VTB5003_TN5003
Class: 0, BENIGN
Pmalign: 0.013
PRS: 1.163
Transverse
GradCAM benign
GradCAM malign
Longitudinal
GradCAM benign
GradCAM malign
ID: VTB5006_TN5006
Class: 0, BENIGN
Pmalign: 0.027
PRS: -2.039
Transverse
GradCAM benign
GradCAM malign
Longitudinal
GradCAM benign
GradCAM malign

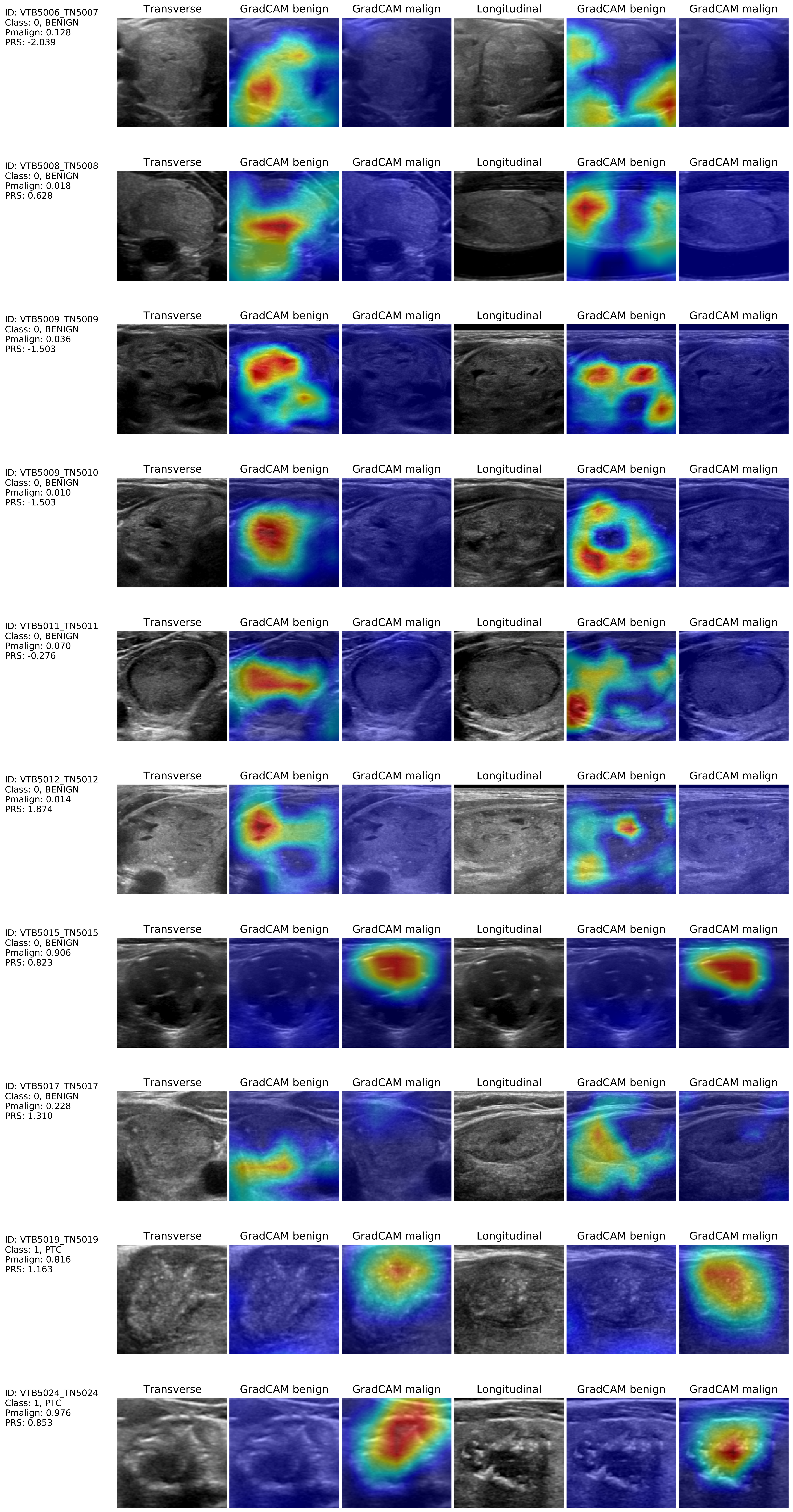

ID: VTB5006_TN5007
Class: 0, BENIGN
Pmalign: 0.128
PRS: -2.039
Transverse
GradCAM benign
GradCAM malign
Longitudinal
GradCAM benign
GradCAM malign
ID: VTB5008_TN5008
Class: 0, BENIGN
Pmalign: 0.018
PRS: 0.628
Transverse
GradCAM benign
GradCAM malign
Longitudinal
GradCAM benign
GradCAM malign
ID: VTB5009_TN5009
Class: 0, BENIGN
Pmalign: 0.036
PRS: -1.503
Transverse
GradCAM benign
GradCAM malign
Longitudinal
GradCAM benign
GradCAM malign
ID: VTB5009_TN5010
Class: 0, BENIGN
Pmalign: 0.010
PRS: -1.503
Transverse
GradCAM benign
GradCAM malign
Longitudinal
GradCAM benign
GradCAM malign
ID: VTB5011_TN5011
Class: 0, BENIGN
Pmalign: 0.070
PRS: -0.276
Transverse
GradCAM benign
GradCAM malign
Longitudinal
GradCAM benign
GradCAM malign
ID: VTB5012_TN5012
Class: 0, BENIGN
Pmalign: 0.014
PRS: 1.874
Transverse
GradCAM benign
GradCAM malign
Longitudinal
GradCAM benign
GradCAM malign
ID: VTB5015_TN5015
Class: 0, BENIGN
Pmalign: 0.906
PRS: 0.823
Transverse
GradCAM benign
GradCAM malign
Longitudinal
GradCAM benign
GradCAM malign
ID: VTB5017_TN5017
Class: 0, BENIGN
Pmalign: 0.228
PRS: 1.310
Transverse
GradCAM benign
GradCAM malign
Longitudinal
GradCAM benign
GradCAM malign
ID: VTB5019_TN5019
Class: 1, PTC
Pmalign: 0.816
PRS: 1.163
Transverse
GradCAM benign
GradCAM malign
Longitudinal
GradCAM benign
GradCAM malign
ID: VTB5024_TN5024
Class: 1, PTC
Pmalign: 0.976
PRS: 0.853
Transverse
GradCAM benign
GradCAM malign
Longitudinal
GradCAM benign
GradCAM malign

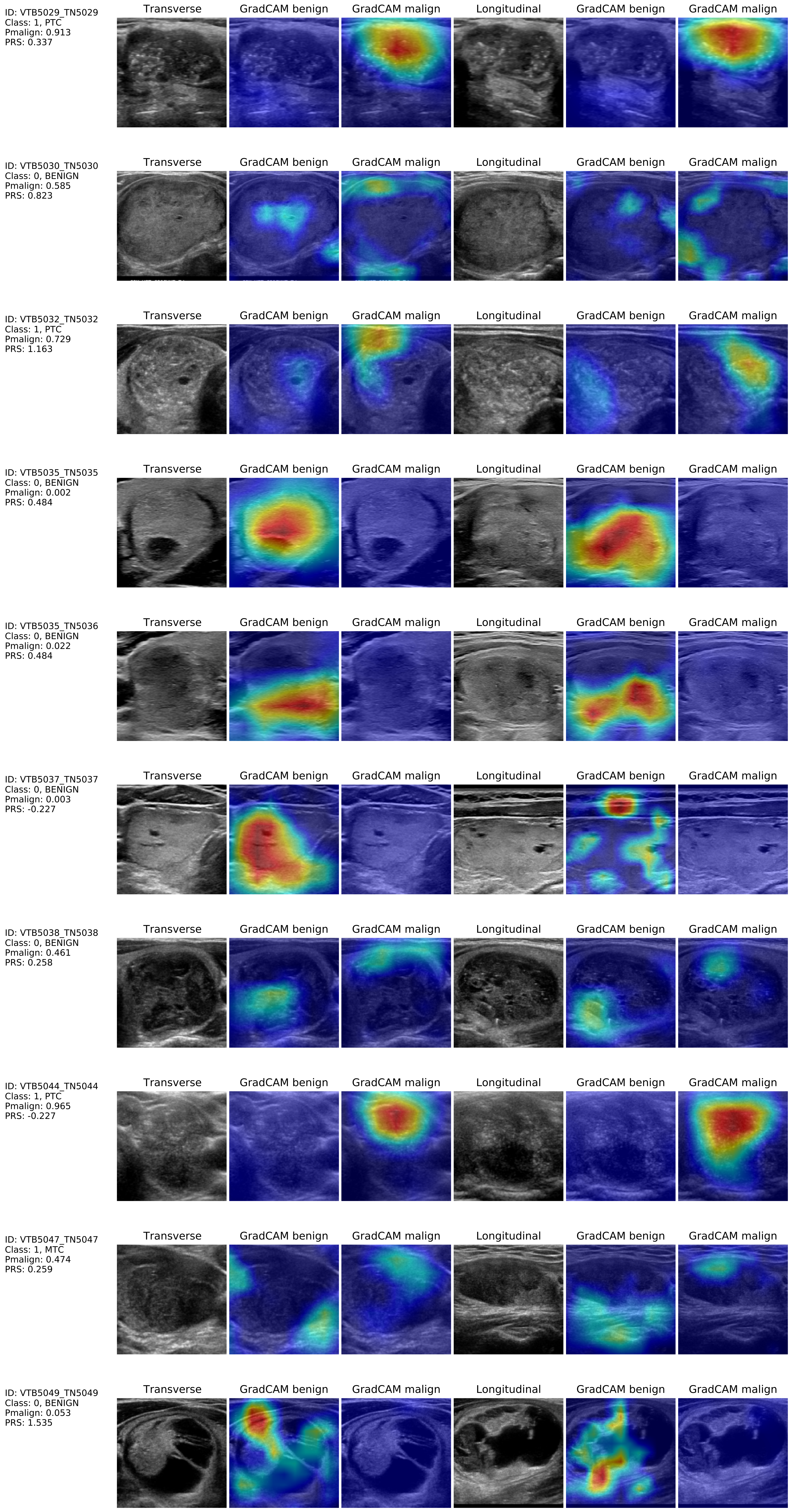
ID: VTB5029_TN5029
Class: 1, PTC
Pmalign: 0.913
PRS: 0.337
Transverse
GradCAM benign
GradCAM malign
Longitudinal
GradCAM benign
GradCAM malign
ID: VTB5030_TN5030
Class: 0, BENIGN
Pmalign: 0.585
PRS: 0.823
Transverse
GradCAM benign
GradCAM malign
Longitudinal
GradCAM benign
GradCAM malign
ID: VTB5032_TN5032
Class: 1, PTC
Pmalign: 0.729
PRS: 1.163
Transverse
GradCAM benign
GradCAM malign
Longitudinal
GradCAM benign
GradCAM malign
ID: VTB5035_TN5035
Class: 0, BENIGN
Pmalign: 0.002
PRS: 0.484
Transverse
GradCAM benign
GradCAM malign
Longitudinal
GradCAM benign
GradCAM malign
ID: VTB5035_TN5036
Class: 0, BENIGN
Pmalign: 0.022
PRS: 0.484
Transverse
GradCAM benign
GradCAM malign
Longitudinal
GradCAM benign
GradCAM malign
ID: VTB5037_TN5037
Class: 0, BENIGN
Pmalign: 0.003
PRS: -0.227
Transverse
GradCAM benign
GradCAM malign
Longitudinal
GradCAM benign
GradCAM malign
ID: VTB5038_TN5038
Class: 0, BENIGN
Pmalign: 0.461
PRS: 0.258
Transverse
GradCAM benign
GradCAM malign
Longitudinal
GradCAM benign
GradCAM malign
ID: VTB5044_TN5044
Class: 1, PTC
Pmalign: 0.965
PRS: -0.227
Transverse
GradCAM benign
GradCAM malign
Longitudinal
GradCAM benign
GradCAM malign
ID: VTB5047_TN5047
Class: 1, MTC
Pmalign: 0.474
PRS: 0.259
Transverse
GradCAM benign
GradCAM malign
Longitudinal
GradCAM benign
GradCAM malign
ID: VTB5049_TN5049
Class: 0, BENIGN
Pmalign: 0.053
PRS: 1.535
Transverse
GradCAM benign
GradCAM malign
Longitudinal
GradCAM benign
GradCAM malign

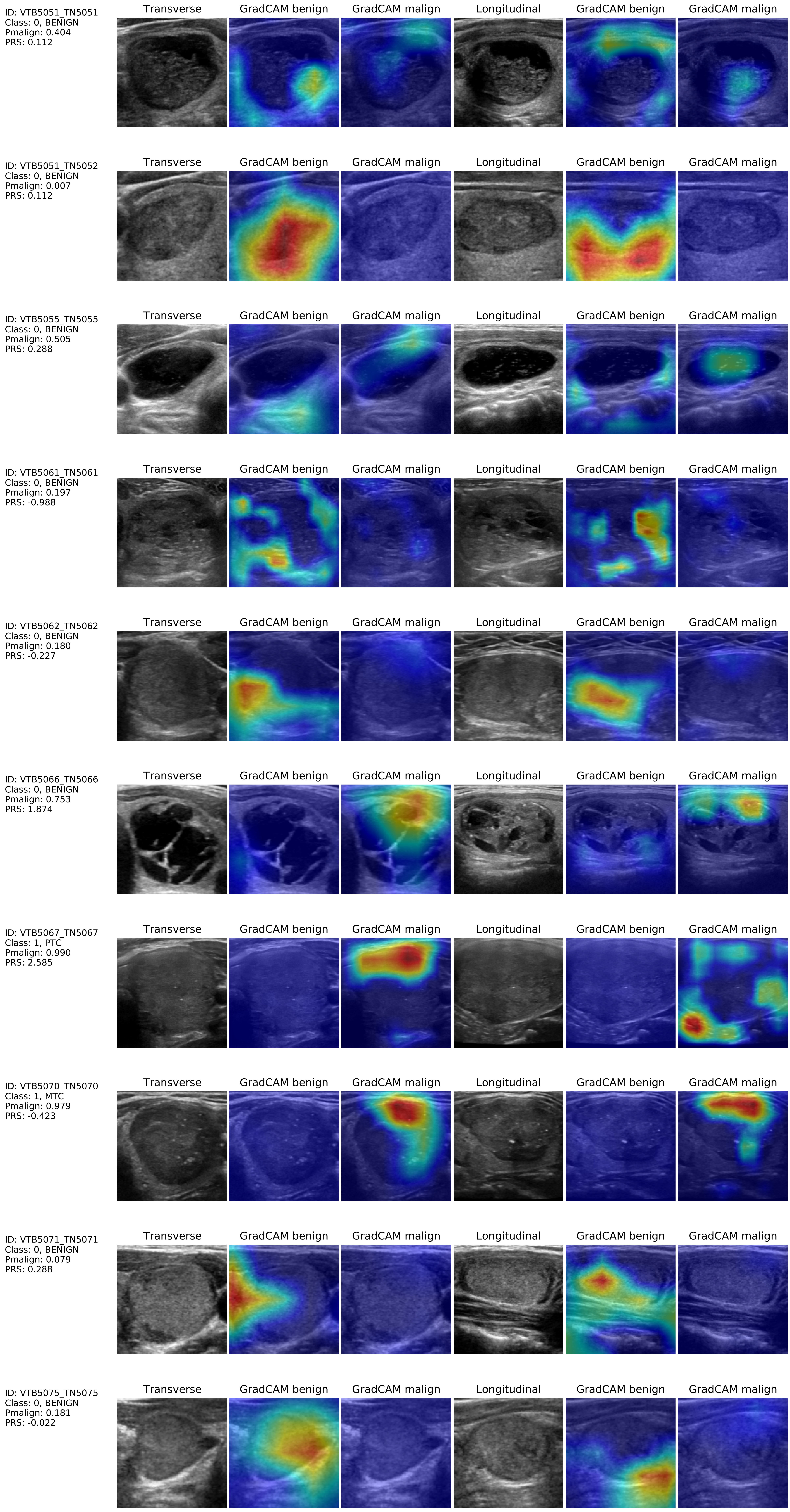

ID: VTB5051_TN5051
Class: 0, BENIGN
Pmalign: 0.404
PRS: 0.112
ID: VTB5051_TN5052
Class: 0, BENIGN
Pmalign: 0.007
PRS: 0.112
ID: VTB5055_TN5055
Class: 0, BENIGN
Pmalign: 0.505
PRS: 0.288
ID: VTB5061_TN5061
Class: 0, BENIGN
Pmalign: 0.197
PRS: -0.988
ID: VTB5062_TN5062
Class: 0, BENIGN
Pmalign: 0.180
PRS: -0.227
ID: VTB5066_TN5066
Class: 0, BENIGN
Pmalign: 0.753
PRS: 1.874
ID: VTB5067_TN5067
Class: 1, PTC
Pmalign: 0.990
PRS: 2.585
ID: VTB5070_TN5070
Class: 1, MTC
Pmalign: 0.979
PRS: -0.423
ID: VTB5071_TN5071
Class: 0, BENIGN
Pmalign: 0.079
PRS: 0.288
ID: VTB5075_TN5075
Class: 0, BENIGN
Pmalign: 0.181
PRS: -0.022
Transverse
GradCAM benign
GradCAM malign
Longitudinal
GradCAM benign
GradCAM malign

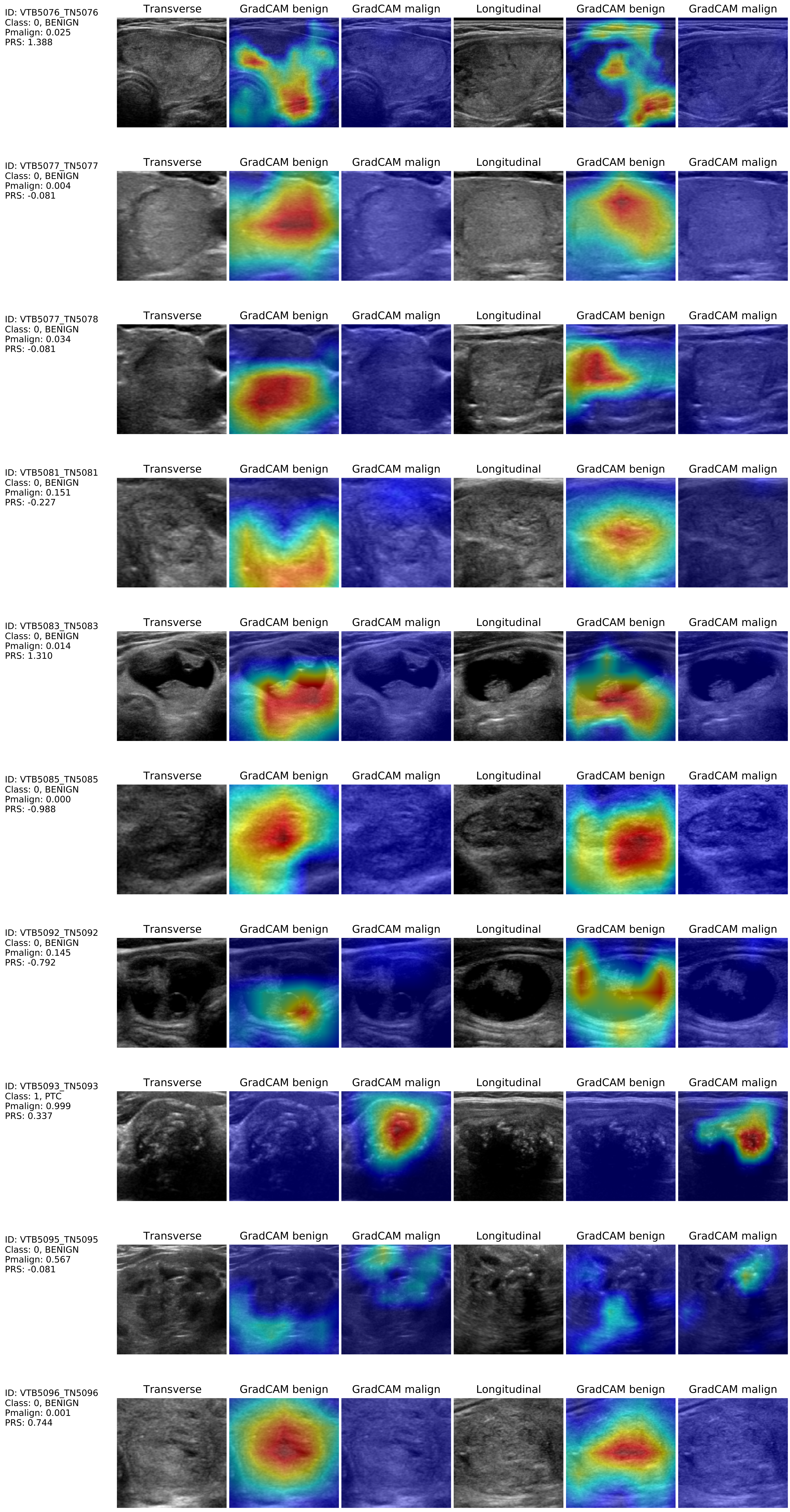
ID: VTB5076_TN5076
Class: 0, BENIGN
Pmalign: 0.025
PRS: 1.388
ID: VTB5077_TN5077
Class: 0, BENIGN
Pmalign: 0.004
PRS: -0.081
ID: VTB5077_TN5078
Class: 0, BENIGN
Pmalign: 0.034
PRS: -0.081
ID: VTB5081_TN5081
Class: 0, BENIGN
Pmalign: 0.151
PRS: -0.227
ID: VTB5083_TN5083
Class: 0, BENIGN
Pmalign: 0.014
PRS: 1.310
ID: VTB5085_TN5085
Class: 0, BENIGN
Pmalign: 0.000
PRS: -0.988
ID: VTB5092_TN5092
Class: 0, BENIGN
Pmalign: 0.145
PRS: -0.792
ID: VTB5093_TN5093
Class: 1, PTC
Pmalign: 0.999
PRS: 0.337
ID: VTB5095_TN5095
Class: 0, BENIGN
Pmalign: 0.567
PRS: -0.081
ID: VTB5096_TN5096
Class: 0, BENIGN
Pmalign: 0.001
PRS: 0.744
Transverse
GradCAM benign
GradCAM malign
Longitudinal
GradCAM benign
GradCAM malign

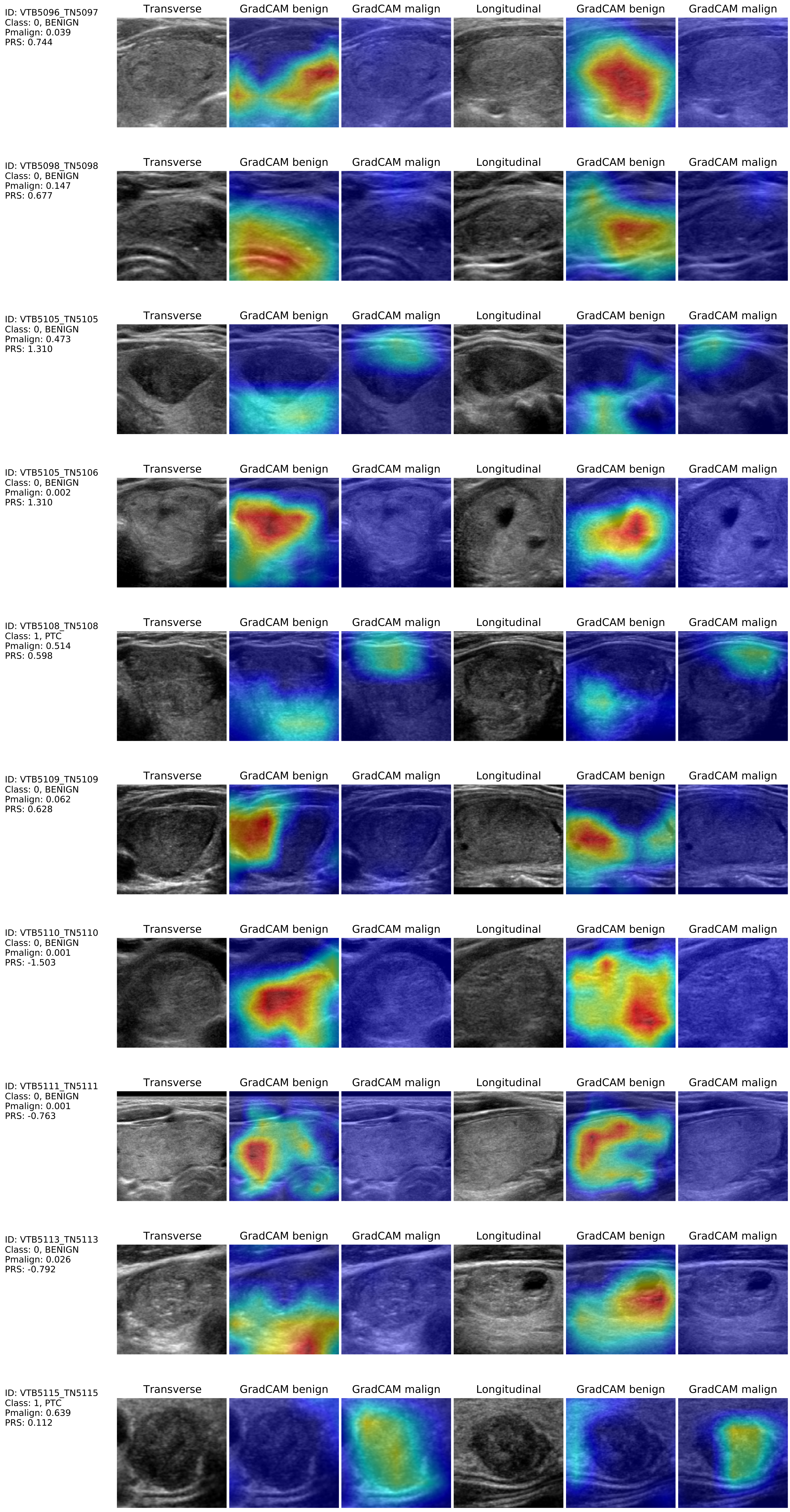
ID: VTB5096_TN5097
Class: 0, BENIGN
Pmalign: 0.039
PRS: 0.744
Transverse
GradCAM benign
GradCAM malign
Longitudinal
GradCAM benign
GradCAM malign
ID: VTB5098_TN5098
Class: 0, BENIGN
Pmalign: 0.147
PRS: 0.677
Transverse
GradCAM benign
GradCAM malign
Longitudinal
GradCAM benign
GradCAM malign
ID: VTB5105_TN5105
Class: 0, BENIGN
Pmalign: 0.473
PRS: 1.310
Transverse
GradCAM benign
GradCAM malign
Longitudinal
GradCAM benign
GradCAM malign
ID: VTB5105_TN5106
Class: 0, BENIGN
Pmalign: 0.002
PRS: 1.310
Transverse
GradCAM benign
GradCAM malign
Longitudinal
GradCAM benign
GradCAM malign
ID: VTB5108_TN5108
Class: 1, PTC
Pmalign: 0.514
PRS: 0.598
Transverse
GradCAM benign
GradCAM malign
Longitudinal
GradCAM benign
GradCAM malign
ID: VTB5109_TN5109
Class: 0, BENIGN
Pmalign: 0.062
PRS: 0.628
Transverse
GradCAM benign
GradCAM malign
Longitudinal
GradCAM benign
GradCAM malign
ID: VTB5110_TN5110
Class: 0, BENIGN
Pmalign: 0.001
PRS: -1.503
Transverse
GradCAM benign
GradCAM malign
Longitudinal
GradCAM benign
GradCAM malign
ID: VTB5111_TN5111
Class: 0, BENIGN
Pmalign: 0.001
PRS: -0.763
Transverse
GradCAM benign
GradCAM malign
Longitudinal
GradCAM benign
GradCAM malign
ID: VTB5113_TN5113
Class: 0, BENIGN
Pmalign: 0.026
PRS: -0.792
Transverse
GradCAM benign
GradCAM malign
Longitudinal
GradCAM benign
GradCAM malign
ID: VTB5115_TN5115
Class: 1, PTC
Pmalign: 0.639
PRS: 0.112
Transverse
GradCAM benign
GradCAM malign
Longitudinal
GradCAM benign
GradCAM malign

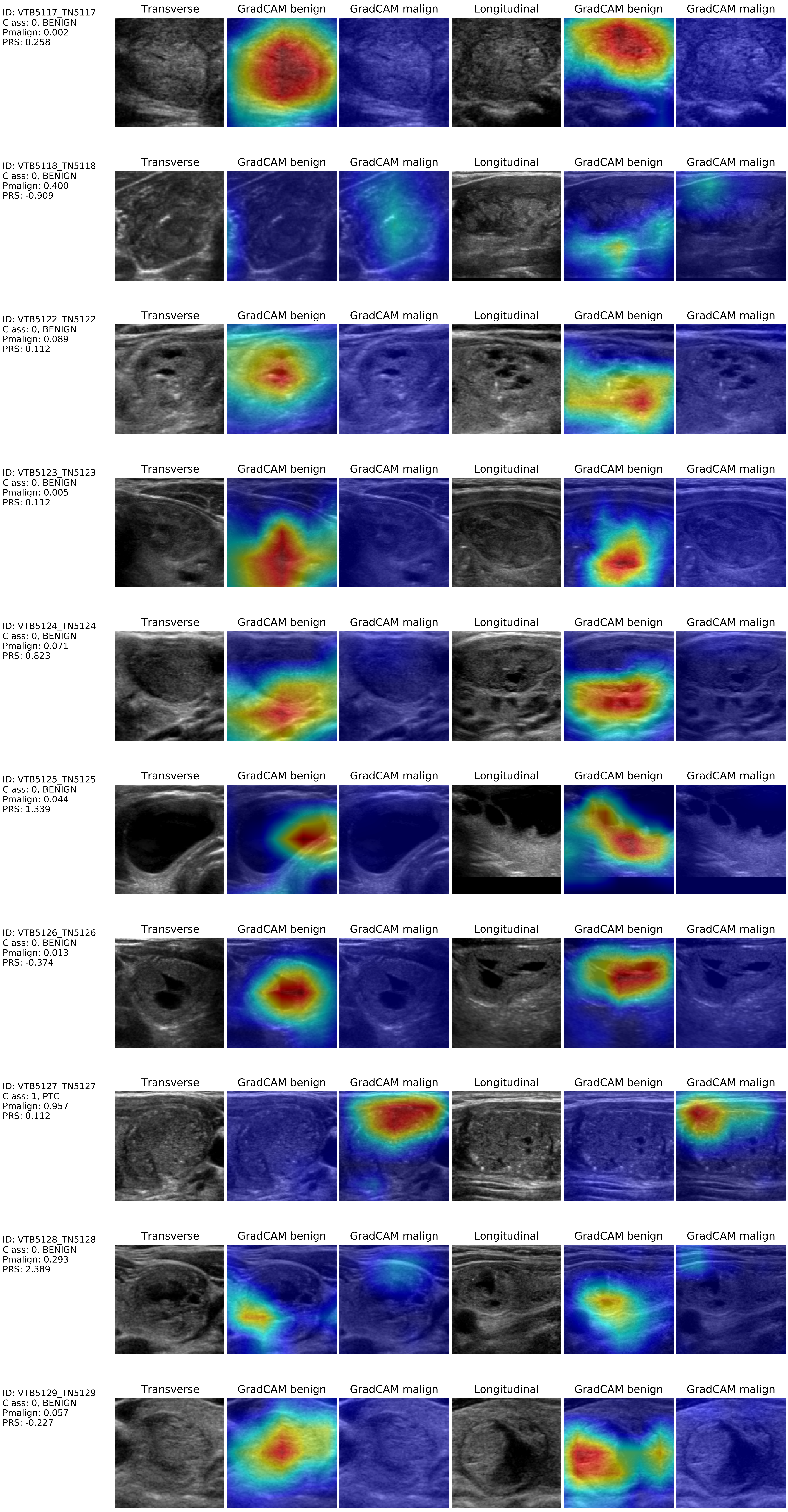
ID: VTB5117_TN5117
Class: 0, BENIGN
Pmalign: 0.002
PRS: 0.258
ID: VTB5118_TN5118
Class: 0, BENIGN
Pmalign: 0.400
PRS: -0.909
ID: VTB5122_TN5122
Class: 0, BENIGN
Pmalign: 0.089
PRS: 0.112
ID: VTB5123_TN5123
Class: 0, BENIGN
Pmalign: 0.005
PRS: 0.112
ID: VTB5124_TN5124
Class: 0, BENIGN
Pmalign: 0.071
PRS: 0.823
ID: VTB5125_TN5125
Class: 0, BENIGN
Pmalign: 0.044
PRS: 1.339
ID: VTB5126_TN5126
Class: 0, BENIGN
Pmalign: 0.013
PRS: -0.374
ID: VTB5127_TN5127
Class: 1, PTC
Pmalign: 0.957
PRS: 0.112
ID: VTB5128_TN5128
Class: 0, BENIGN
Pmalign: 0.293
PRS: 2.389
ID: VTB5129_TN5129
Class: 0, BENIGN
Pmalign: 0.057
PRS: -0.227
Transverse
GradCAM benign
GradCAM malign
Longitudinal
GradCAM benign
GradCAM malign

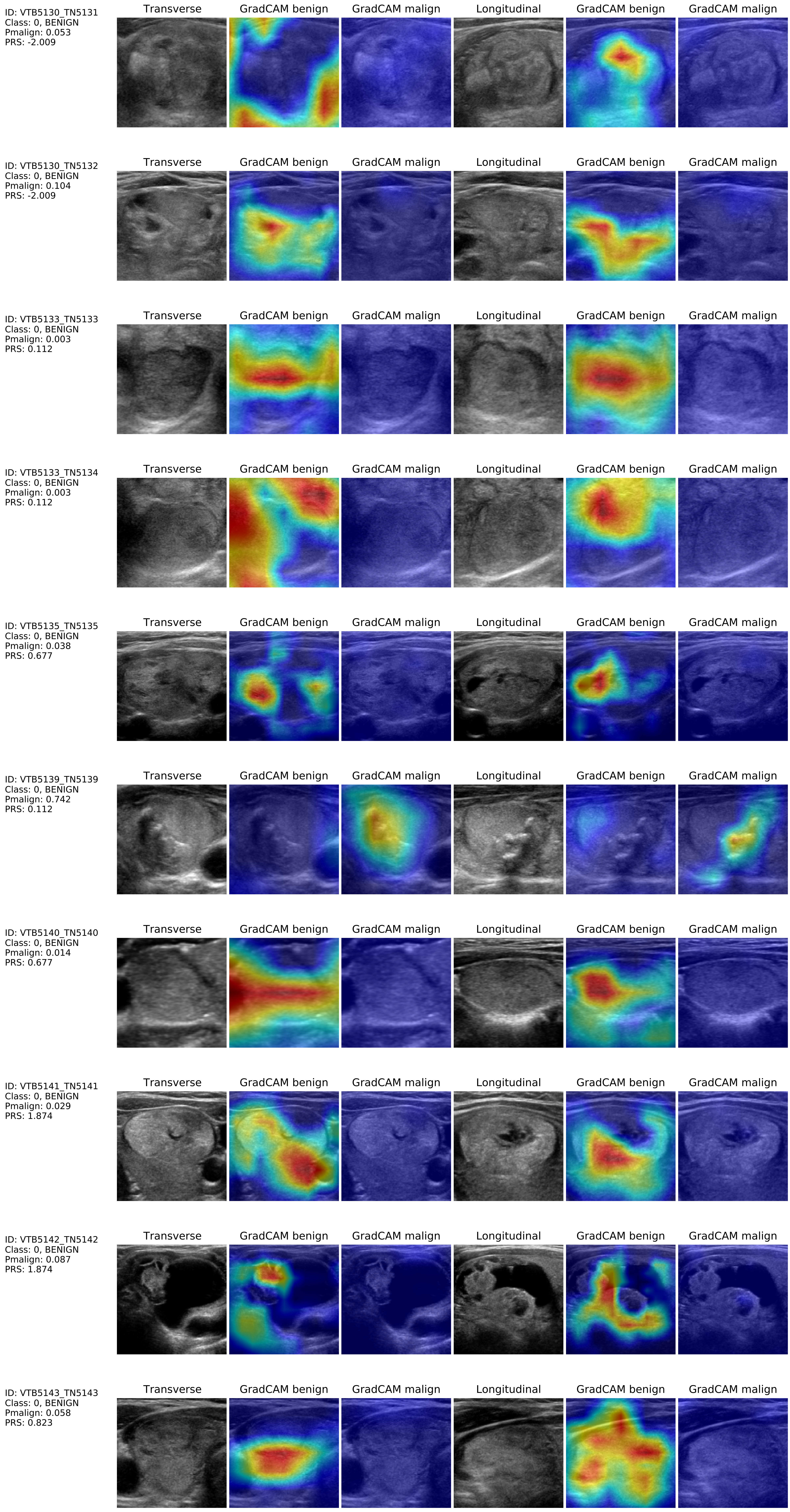
ID: VTB5130_TN5131
Class: 0, BENIGN
Pmalign: 0.053
PRS: -2.009
Transverse
GradCAM benign
GradCAM malign
Longitudinal
GradCAM benign
GradCAM malign
ID: VTB5130_TN5132
Class: 0, BENIGN
Pmalign: 0.104
PRS: -2.009
Transverse
GradCAM benign
GradCAM malign
Longitudinal
GradCAM benign
GradCAM malign
ID: VTB5133_TN5133
Class: 0, BENIGN
Pmalign: 0.003
PRS: 0.112
Transverse
GradCAM benign
GradCAM malign
Longitudinal
GradCAM benign
GradCAM malign
ID: VTB5133_TN5134
Class: 0, BENIGN
Pmalign: 0.003
PRS: 0.112
Transverse
GradCAM benign
GradCAM malign
Longitudinal
GradCAM benign
GradCAM malign
ID: VTB5135_TN5135
Class: 0, BENIGN
Pmalign: 0.038
PRS: 0.677
Transverse
GradCAM benign
GradCAM malign
Longitudinal
GradCAM benign
GradCAM malign
ID: VTB5139_TN5139
Class: 0, BENIGN
Pmalign: 0.742
PRS: 0.112
Transverse
GradCAM benign
GradCAM malign
Longitudinal
GradCAM benign
GradCAM malign
ID: VTB5140_TN5140
Class: 0, BENIGN
Pmalign: 0.014
PRS: 0.677
Transverse
GradCAM benign
GradCAM malign
Longitudinal
GradCAM benign
GradCAM malign
ID: VTB5141_TN5141
Class: 0, BENIGN
Pmalign: 0.029
PRS: 1.874
Transverse
GradCAM benign
GradCAM malign
Longitudinal
GradCAM benign
GradCAM malign
ID: VTB5142_TN5142
Class: 0, BENIGN
Pmalign: 0.087
PRS: 1.874
Transverse
GradCAM benign
GradCAM malign
Longitudinal
GradCAM benign
GradCAM malign
ID: VTB5143_TN5143
Class: 0, BENIGN
Pmalign: 0.058
PRS: 0.823
Transverse
GradCAM benign
GradCAM malign
Longitudinal
GradCAM benign
GradCAM malign

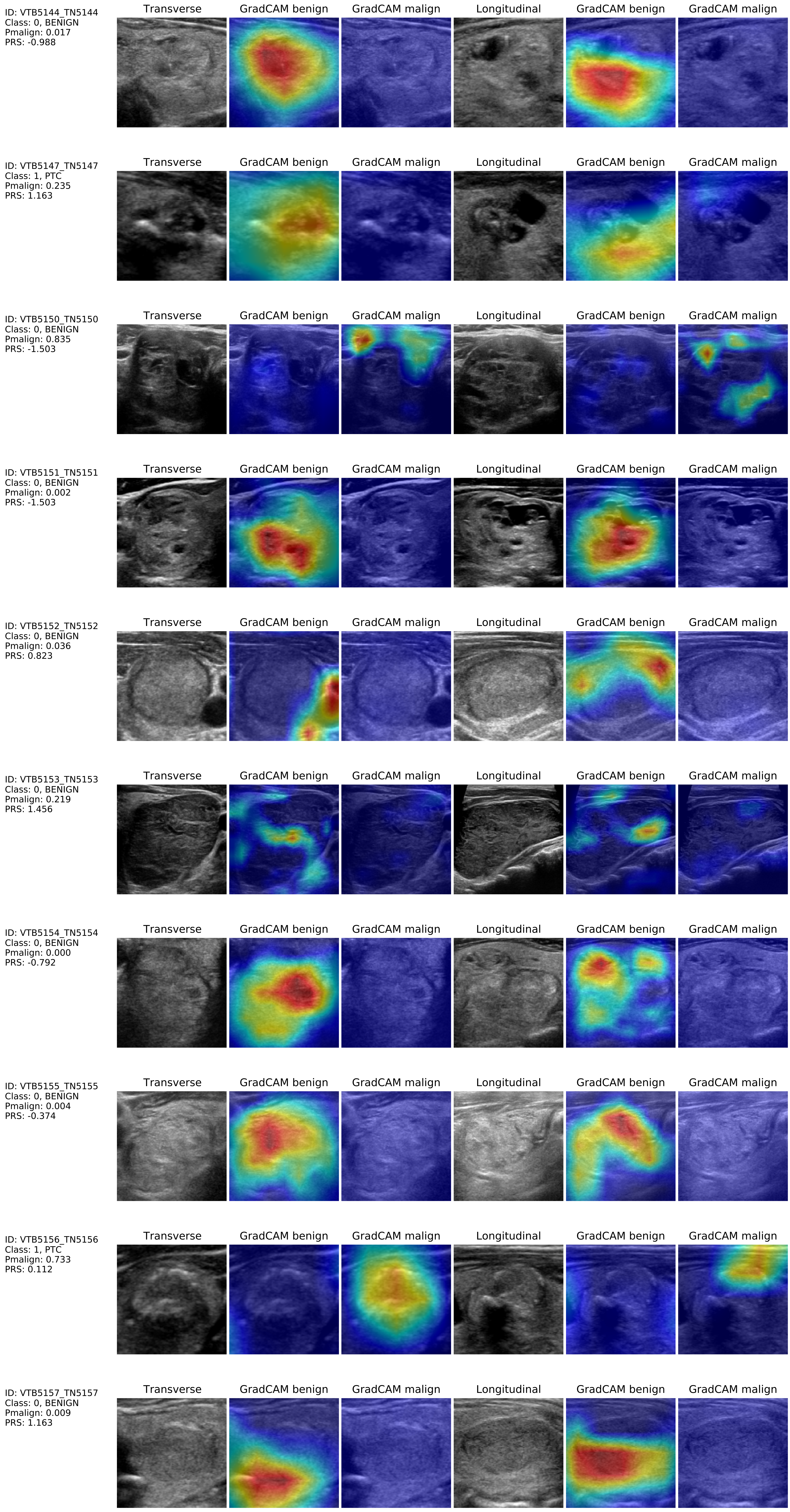

ID: VTB5144_TN5144
Class: 0, BENIGN
Pmalign: 0.017
PRS: -0.988
Transverse
GradCAM benign
GradCAM malign
Longitudinal
GradCAM benign
GradCAM malign
ID: VTB5147_TN5147
Class: 1, PTC
Pmalign: 0.235
PRS: 1.163
Transverse
GradCAM benign
GradCAM malign
Longitudinal
GradCAM benign
GradCAM malign
ID: VTB5150_TN5150
Class: 0, BENIGN
Pmalign: 0.835
PRS: -1.503
Transverse
GradCAM benign
GradCAM malign
Longitudinal
GradCAM benign
GradCAM malign
ID: VTB5151_TN5151
Class: 0, BENIGN
Pmalign: 0.002
PRS: -1.503
Transverse
GradCAM benign
GradCAM malign
Longitudinal
GradCAM benign
GradCAM malign
ID: VTB5152_TN5152
Class: 0, BENIGN
Pmalign: 0.036
PRS: 0.823
Transverse
GradCAM benign
GradCAM malign
Longitudinal
GradCAM benign
GradCAM malign
ID: VTB5153_TN5153
Class: 0, BENIGN
Pmalign: 0.219
PRS: 1.456
Transverse
GradCAM benign
GradCAM malign
Longitudinal
GradCAM benign
GradCAM malign
ID: VTB5154_TN5154
Class: 0, BENIGN
Pmalign: 0.000
PRS: -0.792
Transverse
GradCAM benign
GradCAM malign
Longitudinal
GradCAM benign
GradCAM malign
ID: VTB5155_TN5155
Class: 0, BENIGN
Pmalign: 0.004
PRS: -0.374
Transverse
GradCAM benign
GradCAM malign
Longitudinal
GradCAM benign
GradCAM malign
ID: VTB5156_TN5156
Class: 1, PTC
Pmalign: 0.733
PRS: 0.112
Transverse
GradCAM benign
GradCAM malign
Longitudinal
GradCAM benign
GradCAM malign
ID: VTB5157_TN5157
Class: 0, BENIGN
Pmalign: 0.009
PRS: 1.163
Transverse
GradCAM benign
GradCAM malign
Longitudinal
GradCAM benign
GradCAM malign

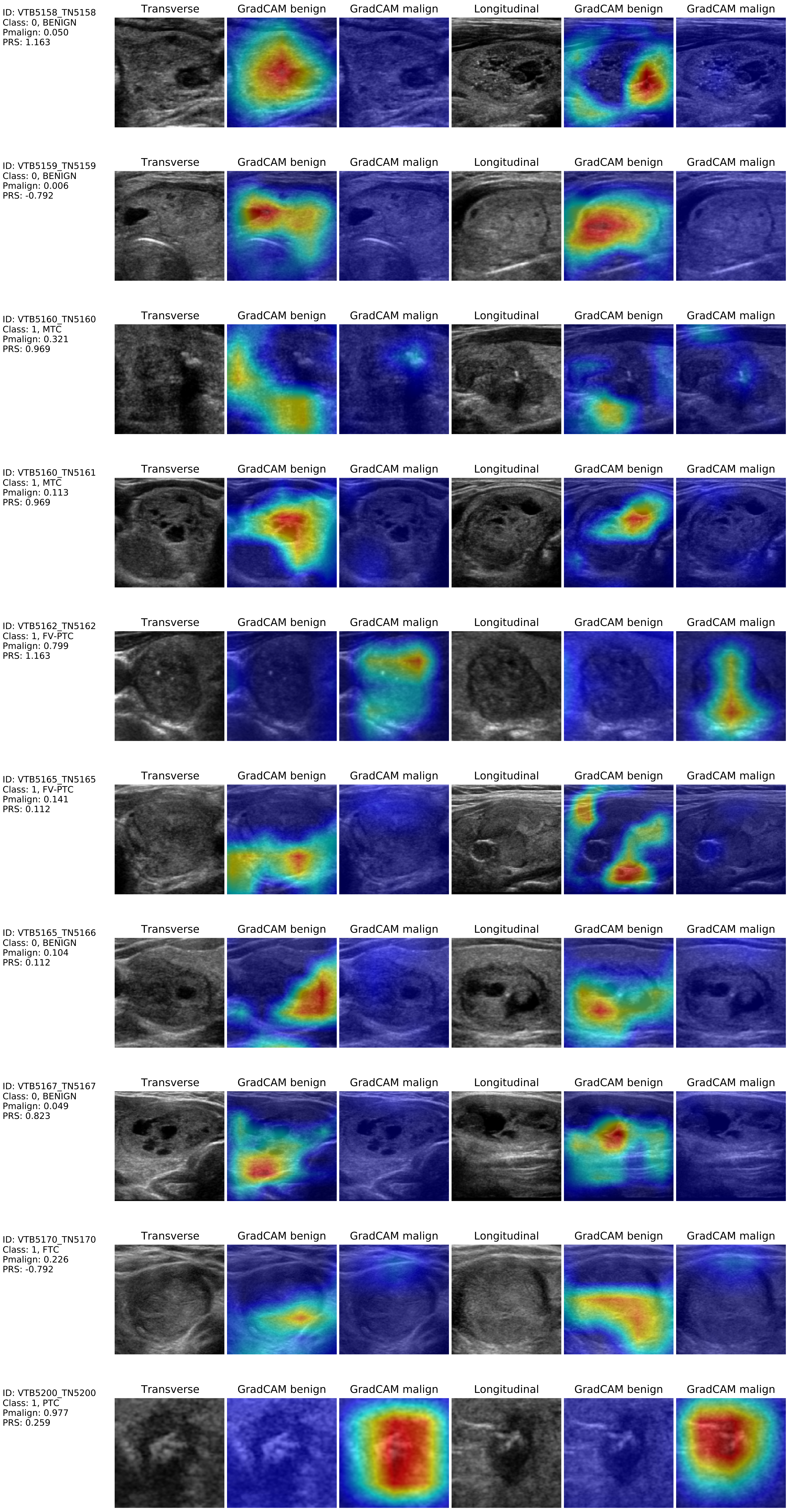
ID: VTB5158_TN5158
Class: 0, BENIGN
Pmalign: 0.050
PRS: 1.163
Transverse
GradCAM benign
GradCAM malign
Longitudinal
GradCAM benign
GradCAM malign
ID: VTB5159_TN5159
Class: 0, BENIGN
Pmalign: 0.006
PRS: -0.792
ID: VTB5160_TN5160
Class: 1, MTC
Pmalign: 0.321
PRS: 0.969
ID: VTB5160_TN5161
Class: 1, MTC
Pmalign: 0.113
PRS: 0.969
ID: VTB5162_TN5162
Class: 1, FV-PTC
Pmalign: 0.799
PRS: 1.163
ID: VTB5165_TN5165
Class: 1, FV-PTC
Pmalign: 0.141
PRS: 0.112
ID: VTB5165_TN5166
Class: 0, BENIGN
Pmalign: 0.104
PRS: 0.112
ID: VTB5167_TN5167
Class: 0, BENIGN
Pmalign: 0.049
PRS: 0.823
ID: VTB5170_TN5170
Class: 1, FTC
Pmalign: 0.226
PRS: -0.792
ID: VTB5200_TN5200
Class: 1, PTC
Pmalign: 0.977
PRS: 0.259

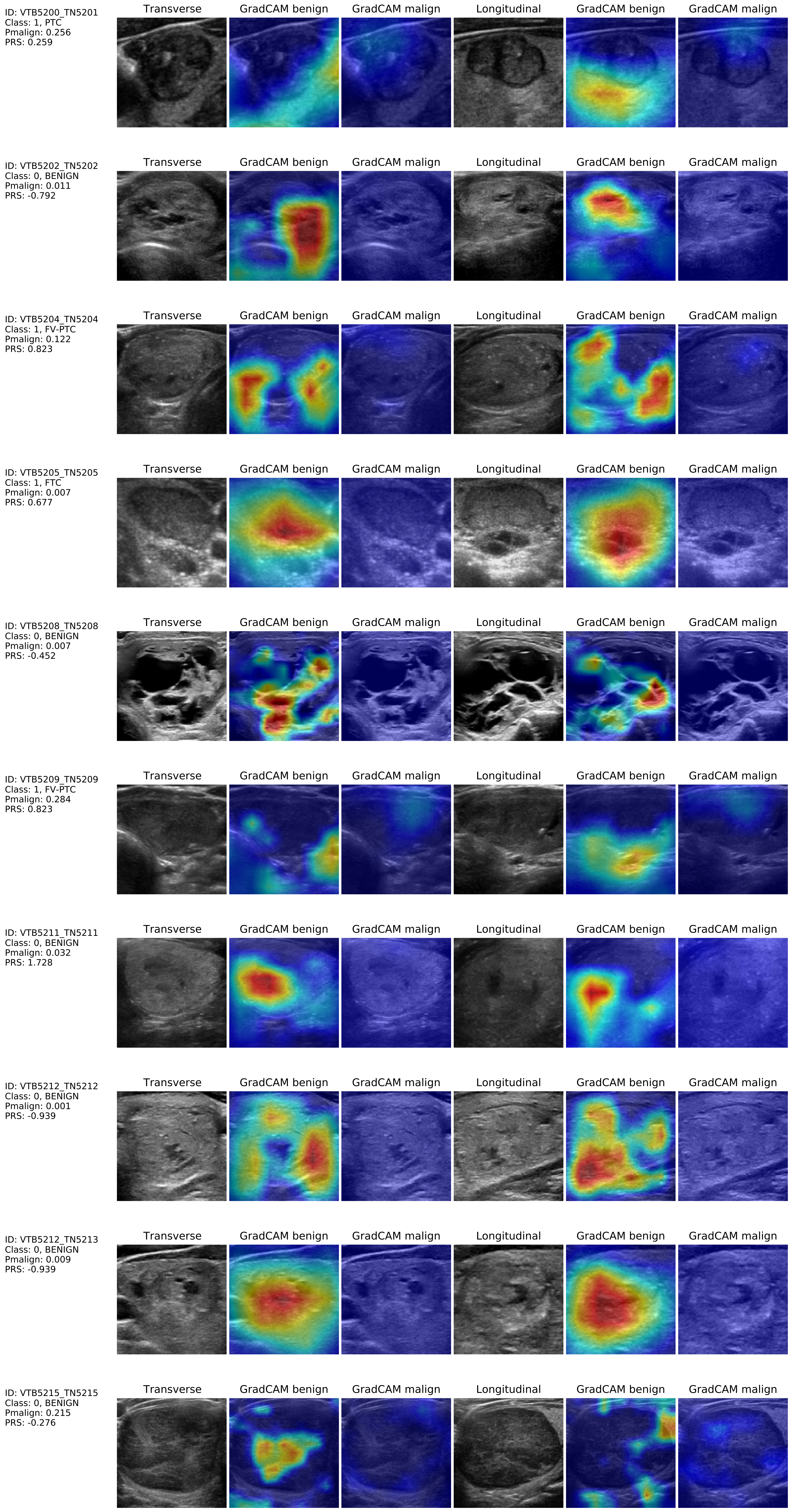

ID: VTB5200_TN5201
Class: 1, PTC
Pmalign: 0.256
PRS: 0.259
Transverse
GradCAM benign
GradCAM malign
Longitudinal
GradCAM benign
GradCAM malign
ID: VTB5202_TN5202
Class: 0, BENIGN
Pmalign: 0.011
PRS: -0.792
Transverse
GradCAM benign
GradCAM malign
Longitudinal
GradCAM benign
GradCAM malign
ID: VTB5204_TN5204
Class: 1, FV-PTC
Pmalign: 0.122
PRS: 0.823
Transverse
GradCAM benign
GradCAM malign
Longitudinal
GradCAM benign
GradCAM malign
ID: VTB5205_TN5205
Class: 1, FTC
Pmalign: 0.007
PRS: 0.677
Transverse
GradCAM benign
GradCAM malign
Longitudinal
GradCAM benign
GradCAM malign
ID: VTB5208_TN5208
Class: 0, BENIGN
Pmalign: 0.007
PRS: -0.452
Transverse
GradCAM benign
GradCAM malign
Longitudinal
GradCAM benign
GradCAM malign
ID: VTB5209_TN5209
Class: 1, FV-PTC
Pmalign: 0.284
PRS: 0.823
Transverse
GradCAM benign
GradCAM malign
Longitudinal
GradCAM benign
GradCAM malign
ID: VTB5211_TN5211
Class: 0, BENIGN
Pmalign: 0.032
PRS: 1.728
Transverse
GradCAM benign
GradCAM malign
Longitudinal
GradCAM benign
GradCAM malign
ID: VTB5212_TN5212
Class: 0, BENIGN
Pmalign: 0.001
PRS: -0.939
Transverse
GradCAM benign
GradCAM malign
Longitudinal
GradCAM benign
GradCAM malign
ID: VTB5212_TN5213
Class: 0, BENIGN
Pmalign: 0.009
PRS: -0.939
Transverse
GradCAM benign
GradCAM malign
Longitudinal
GradCAM benign
GradCAM malign
ID: VTB5215_TN5215
Class: 0, BENIGN
Pmalign: 0.215
PRS: -0.276
Transverse
GradCAM benign
GradCAM malign
Longitudinal
GradCAM benign
GradCAM malign

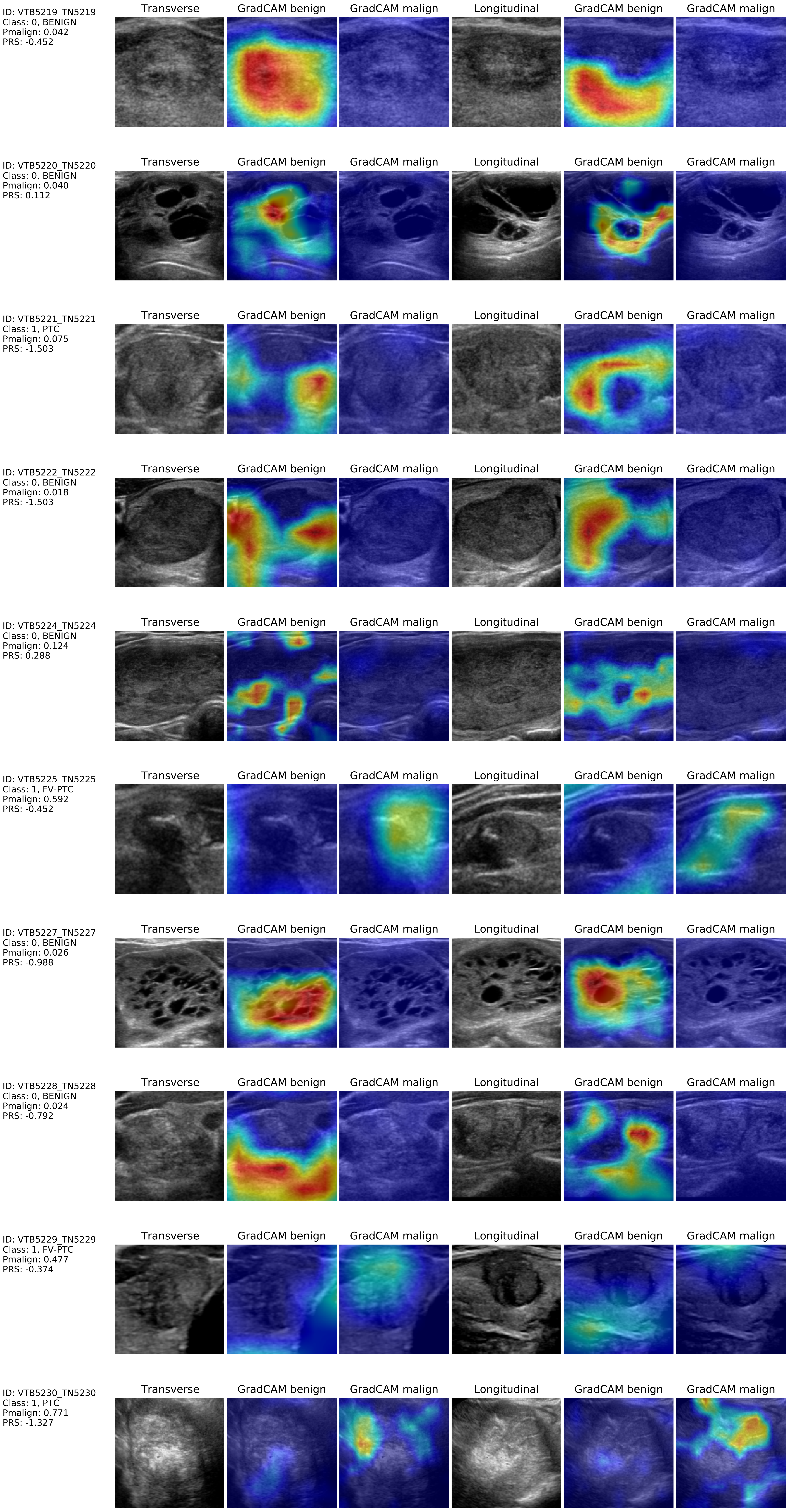
ID: VTB5219_TN5219
Class: 0, BENIGN
Pmalign: 0.042
PRS: -0.452
Transverse
GradCAM benign
GradCAM malign
Longitudinal
GradCAM benign
GradCAM malign
ID: VTB5220_TN5220
Class: 0, BENIGN
Pmalign: 0.040
PRS: 0.112
Transverse
GradCAM benign
GradCAM malign
Longitudinal
GradCAM benign
GradCAM malign
ID: VTB5221_TN5221
Class: 1, PTC
Pmalign: 0.075
PRS: -1.503
Transverse
GradCAM benign
GradCAM malign
Longitudinal
GradCAM benign
GradCAM malign
ID: VTB5222_TN5222
Class: 0, BENIGN
Pmalign: 0.018
PRS: -1.503
Transverse
GradCAM benign
GradCAM malign
Longitudinal
GradCAM benign
GradCAM malign
ID: VTB5224_TN5224
Class: 0, BENIGN
Pmalign: 0.124
PRS: 0.288
Transverse
GradCAM benign
GradCAM malign
Longitudinal
GradCAM benign
GradCAM malign
ID: VTB5225_TN5225
Class: 1, FV-PTC
Pmalign: 0.592
PRS: -0.452
Transverse
GradCAM benign
GradCAM malign
Longitudinal
GradCAM benign
GradCAM malign
ID: VTB5227_TN5227
Class: 0, BENIGN
Pmalign: 0.026
PRS: -0.988
Transverse
GradCAM benign
GradCAM malign
Longitudinal
GradCAM benign
GradCAM malign
ID: VTB5228_TN5228
Class: 0, BENIGN
Pmalign: 0.024
PRS: -0.792
Transverse
GradCAM benign
GradCAM malign
Longitudinal
GradCAM benign
GradCAM malign
ID: VTB5229_TN5229
Class: 1, FV-PTC
Pmalign: 0.477
PRS: -0.374
Transverse
GradCAM benign
GradCAM malign
Longitudinal
GradCAM benign
GradCAM malign
ID: VTB5230_TN5230
Class: 1, PTC
Pmalign: 0.771
PRS: -1.327
Transverse
GradCAM benign
GradCAM malign
Longitudinal
GradCAM benign
GradCAM malign

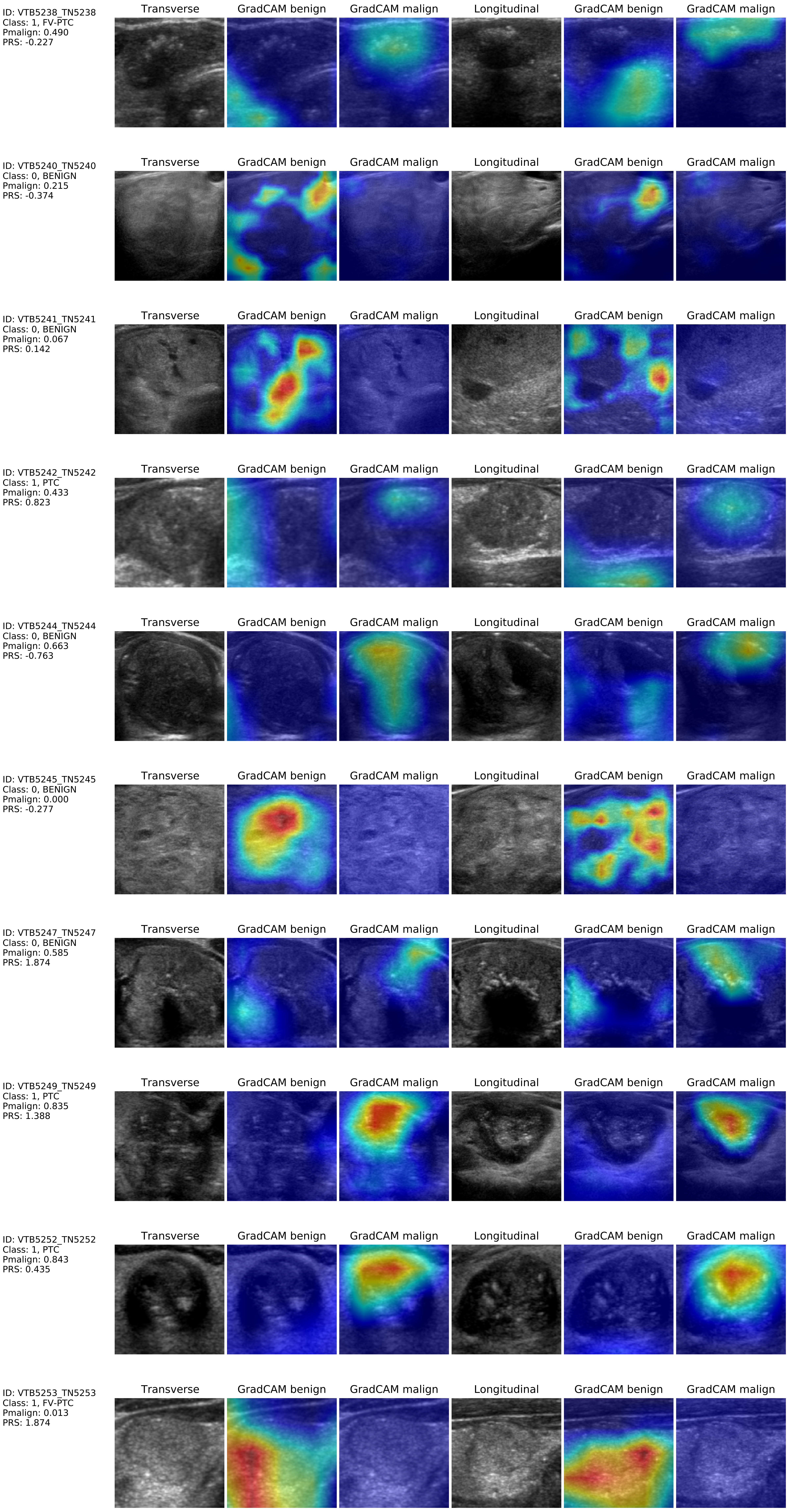
Transverse
GradCAM benign
GradCAM malign
Longitudinal
GradCAM benign
GradCAM malign
ID: VTB5238_TN5238
Class: 1, FV-PTC
Pmalign: 0.490
PRS: -0.227
ID: VTB5240_TN5240
Class: 0, BENIGN
Pmalign: 0.215
PRS: -0.374
ID: VTB5241_TN5241
Class: 0, BENIGN
Pmalign: 0.067
PRS: 0.142
ID: VTB5242_TN5242
Class: 1, PTC
Pmalign: 0.433
PRS: 0.823
ID: VTB5244_TN5244
Class: 0, BENIGN
Pmalign: 0.663
PRS: -0.763
ID: VTB5245_TN5245
Class: 0, BENIGN
Pmalign: 0.000
PRS: -0.277
ID: VTB5247_TN5247
Class: 0, BENIGN
Pmalign: 0.585
PRS: 1.874
ID: VTB5249_TN5249
Class: 1, PTC
Pmalign: 0.835
PRS: 1.388
ID: VTB5252_TN5252
Class: 1, PTC
Pmalign: 0.843
PRS: 0.435
ID: VTB5253_TN5253
Class: 1, FV-PTC
Pmalign: 0.013
PRS: 1.874

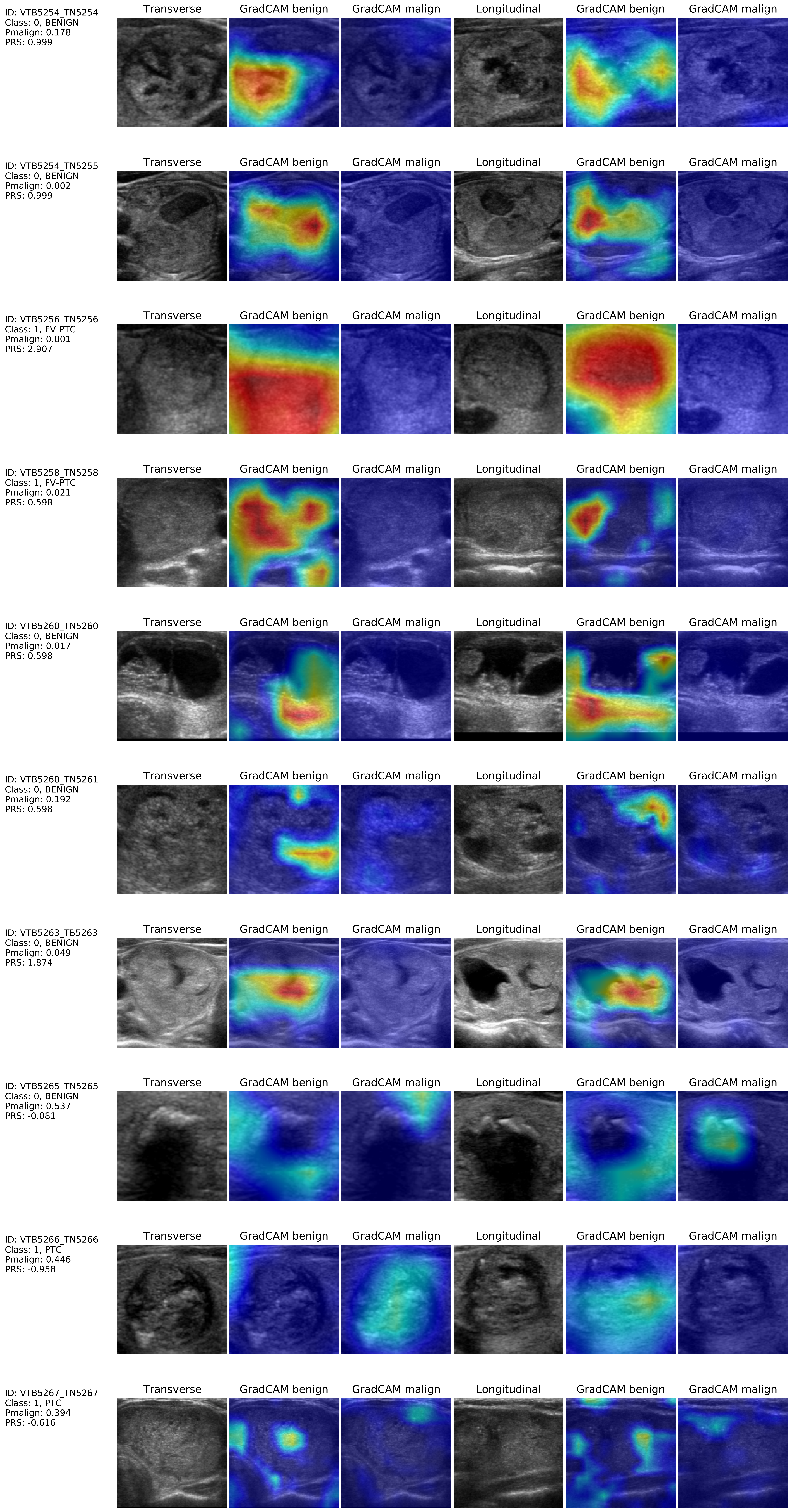
Transverse
GradCAM benign
GradCAM malign
Longitudinal
GradCAM benign
GradCAM malign
ID: VTB5254_TN5254
Class: 0, BENIGN
Pmalign: 0.178
PRS: 0.999
ID: VTB5254_TN5255
Class: 0, BENIGN
Pmalign: 0.002
PRS: 0.999
ID: VTB5256_TN5256
Class: 1, FV-PTC
Pmalign: 0.001
PRS: 2.907
ID: VTB5258_TN5258
Class: 1, FV-PTC
Pmalign: 0.021
PRS: 0.598
ID: VTB5260_TN5260
Class: 0, BENIGN
Pmalign: 0.017
PRS: 0.598
ID: VTB5260_TN5261
Class: 0, BENIGN
Pmalign: 0.192
PRS: 0.598
ID: VTB5263_TB5263
Class: 0, BENIGN
Pmalign: 0.049
PRS: 1.874
ID: VTB5265_TN5265
Class: 0, BENIGN
Pmalign: 0.537
PRS: -0.081
ID: VTB5266_TN5266
Class: 1, PTC
Pmalign: 0.446
PRS: -0.958
ID: VTB5267_TN5267
Class: 1, PTC
Pmalign: 0.394
PRS: -0.616

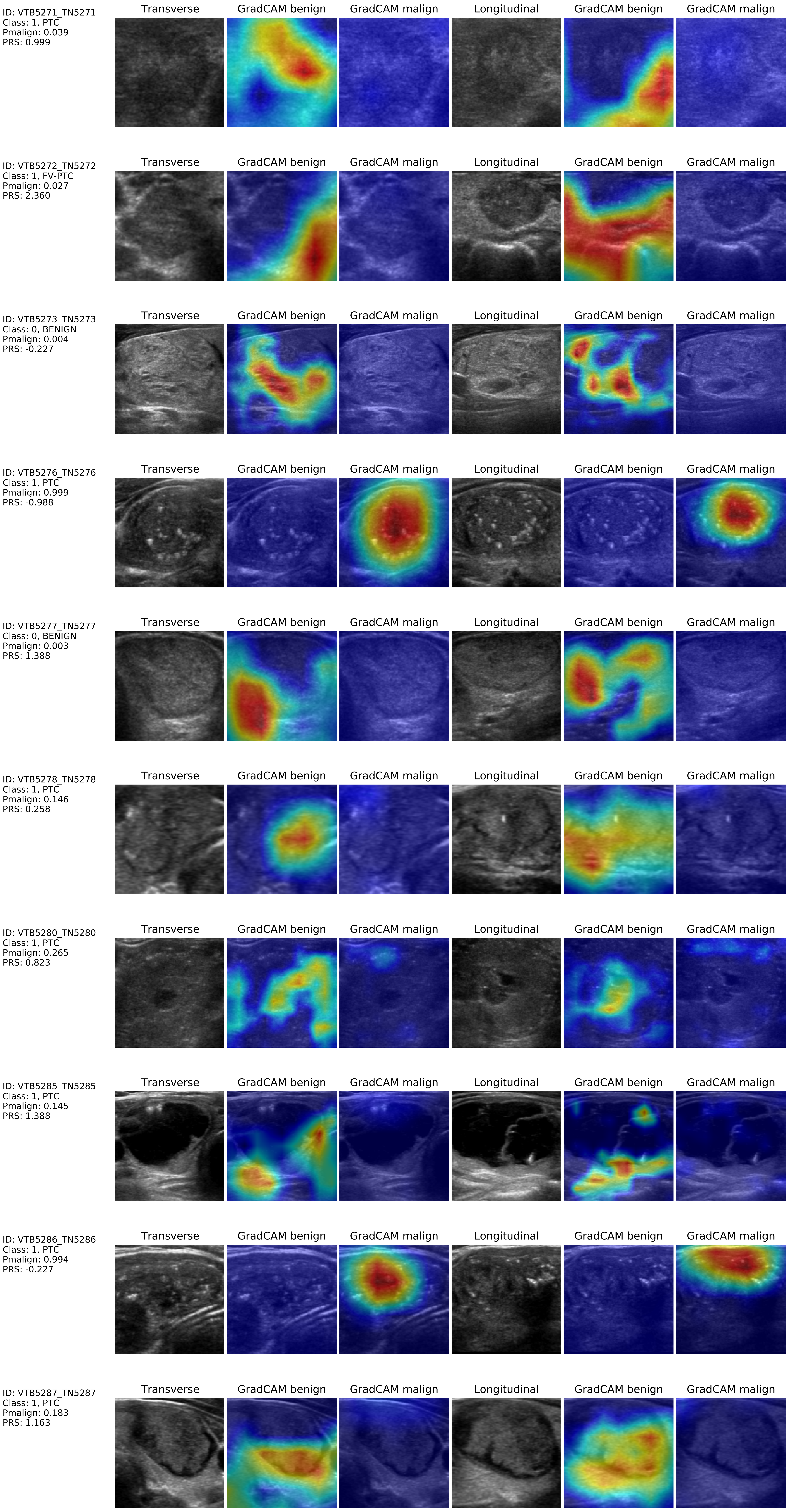
ID: VTB5271_TN5271
Class: 1, PTC
Pmalign: 0.039
PRS: 0.999
Transverse
GradCAM benign
GradCAM malign
Longitudinal
GradCAM benign
GradCAM malign
ID: VTB5272_TN5272
Class: 1, FV-PTC
Pmalign: 0.027
PRS: 2.360
Transverse
GradCAM benign
GradCAM malign
Longitudinal
GradCAM benign
GradCAM malign
ID: VTB5273_TN5273
Class: 0, BENIGN
Pmalign: 0.004
PRS: -0.227
Transverse
GradCAM benign
GradCAM malign
Longitudinal
GradCAM benign
GradCAM malign
ID: VTB5276_TN5276
Class: 1, PTC
Pmalign: 0.999
PRS: -0.988
Transverse
GradCAM benign
GradCAM malign
Longitudinal
GradCAM benign
GradCAM malign
ID: VTB5277_TN5277
Class: 0, BENIGN
Pmalign: 0.003
PRS: 1.388
Transverse
GradCAM benign
GradCAM malign
Longitudinal
GradCAM benign
GradCAM malign
ID: VTB5278_TN5278
Class: 1, PTC
Pmalign: 0.146
PRS: 0.258
Transverse
GradCAM benign
GradCAM malign
Longitudinal
GradCAM benign
GradCAM malign
ID: VTB5280_TN5280
Class: 1, PTC
Pmalign: 0.265
PRS: 0.823
Transverse
GradCAM benign
GradCAM malign
Longitudinal
GradCAM benign
GradCAM malign
ID: VTB5285_TN5285
Class: 1, PTC
Pmalign: 0.145
PRS: 1.388
Transverse
GradCAM benign
GradCAM malign
Longitudinal
GradCAM benign
GradCAM malign
ID: VTB5286_TN5286
Class: 1, PTC
Pmalign: 0.994
PRS: -0.227
Transverse
GradCAM benign
GradCAM malign
Longitudinal
GradCAM benign
GradCAM malign
ID: VTB5287_TN5287
Class: 1, PTC
Pmalign: 0.183
PRS: 1.163
Transverse
GradCAM benign
GradCAM malign
Longitudinal
GradCAM benign
GradCAM malign

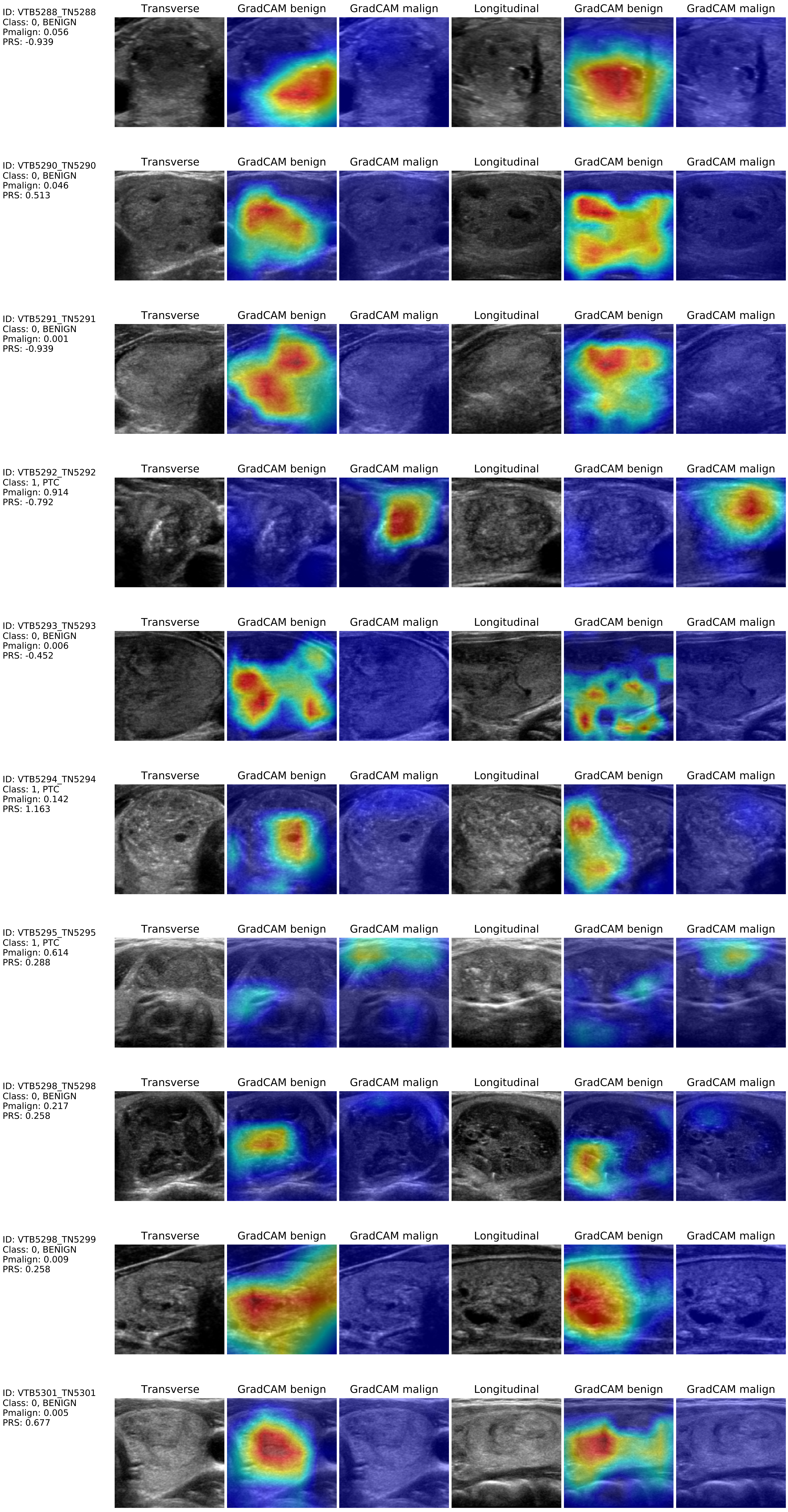
ID: VTB5288_TN5288
Class: 0, BENIGN
Pmalign: 0.056
PRS: -0.939
ID: VTB5290_TN5290
Class: 0, BENIGN
Pmalign: 0.046
PRS: 0.513
ID: VTB5291_TN5291
Class: 0, BENIGN
Pmalign: 0.001
PRS: -0.939
ID: VTB5292_TN5292
Class: 1, PTC
Pmalign: 0.914
PRS: -0.792
ID: VTB5293_TN5293
Class: 0, BENIGN
Pmalign: 0.006
PRS: -0.452
ID: VTB5294_TN5294
Class: 1, PTC
Pmalign: 0.142
PRS: 1.163
ID: VTB5295_TN5295
Class: 1, PTC
Pmalign: 0.614
PRS: 0.288
ID: VTB5298_TN5298
Class: 0, BENIGN
Pmalign: 0.217
PRS: 0.258
ID: VTB5298_TN5299
Class: 0, BENIGN
Pmalign: 0.009
PRS: 0.258
ID: VTB5301_TN5301
Class: 0, BENIGN
Pmalign: 0.005
PRS: 0.677
Transverse
GradCAM benign
GradCAM malign
Longitudinal
GradCAM benign
GradCAM malign

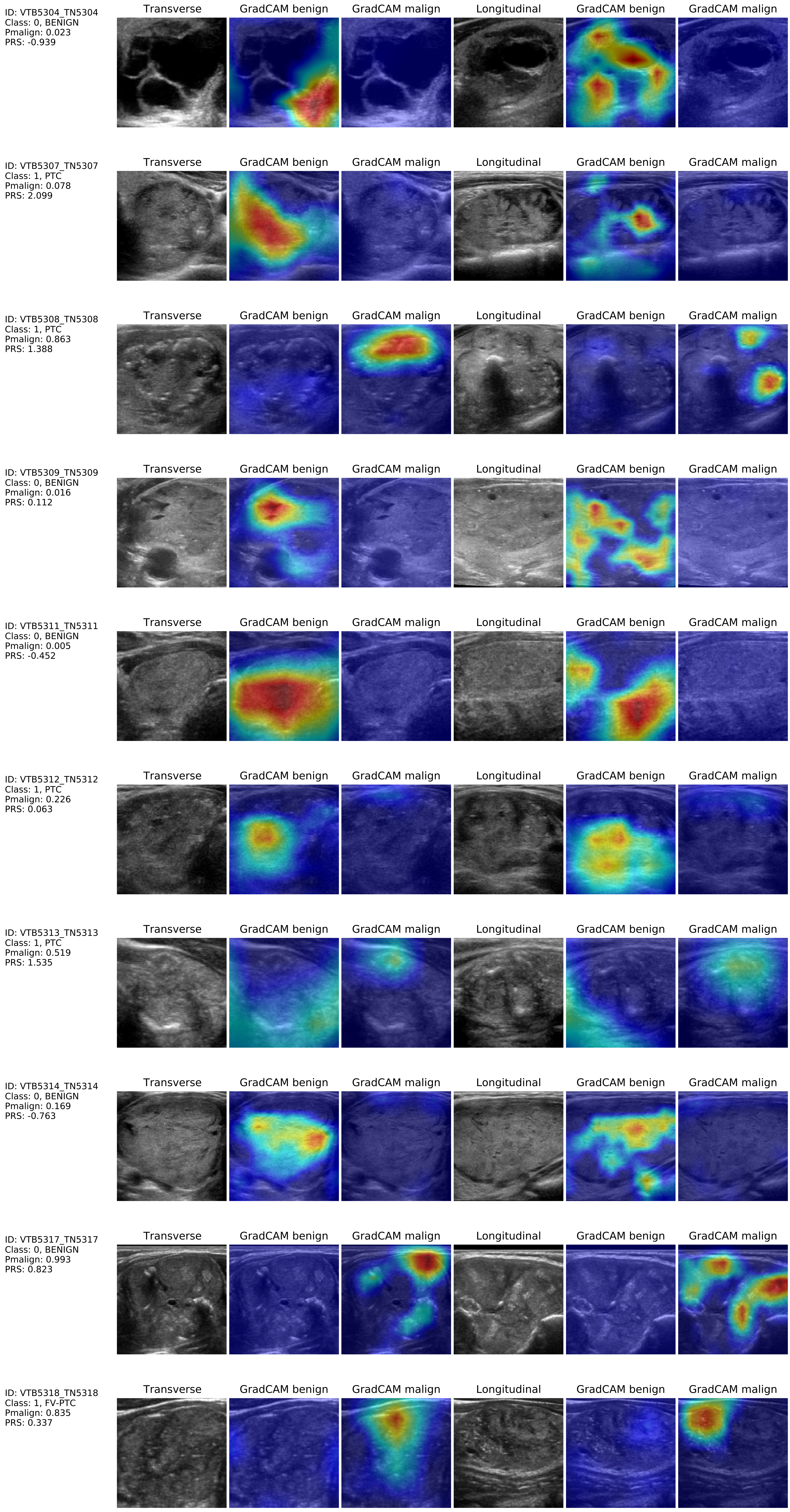
Transverse
GradCAM benign
GradCAM malign
Longitudinal
GradCAM benign
GradCAM malign
ID: VTB5304_TN5304
Class: 0, BENIGN
Pmalign: 0.023
PRS: -0.939
ID: VTB5307_TN5307
Class: 1, PTC
Pmalign: 0.078
PRS: 2.099
ID: VTB5308_TN5308
Class: 1, PTC
Pmalign: 0.863
PRS: 1.388
ID: VTB5309_TN5309
Class: 0, BENIGN
Pmalign: 0.016
PRS: 0.112
ID: VTB5311_TN5311
Class: 0, BENIGN
Pmalign: 0.005
PRS: -0.452
ID: VTB5312_TN5312
Class: 1, PTC
Pmalign: 0.226
PRS: 0.063
ID: VTB5313_TN5313
Class: 1, PTC
Pmalign: 0.519
PRS: 1.535
ID: VTB5314_TN5314
Class: 0, BENIGN
Pmalign: 0.169
PRS: -0.763
ID: VTB5317_TN5317
Class: 0, BENIGN
Pmalign: 0.993
PRS: 0.823
ID: VTB5318_TN5318
Class: 1, FV-PTC
Pmalign: 0.835
PRS: 0.337

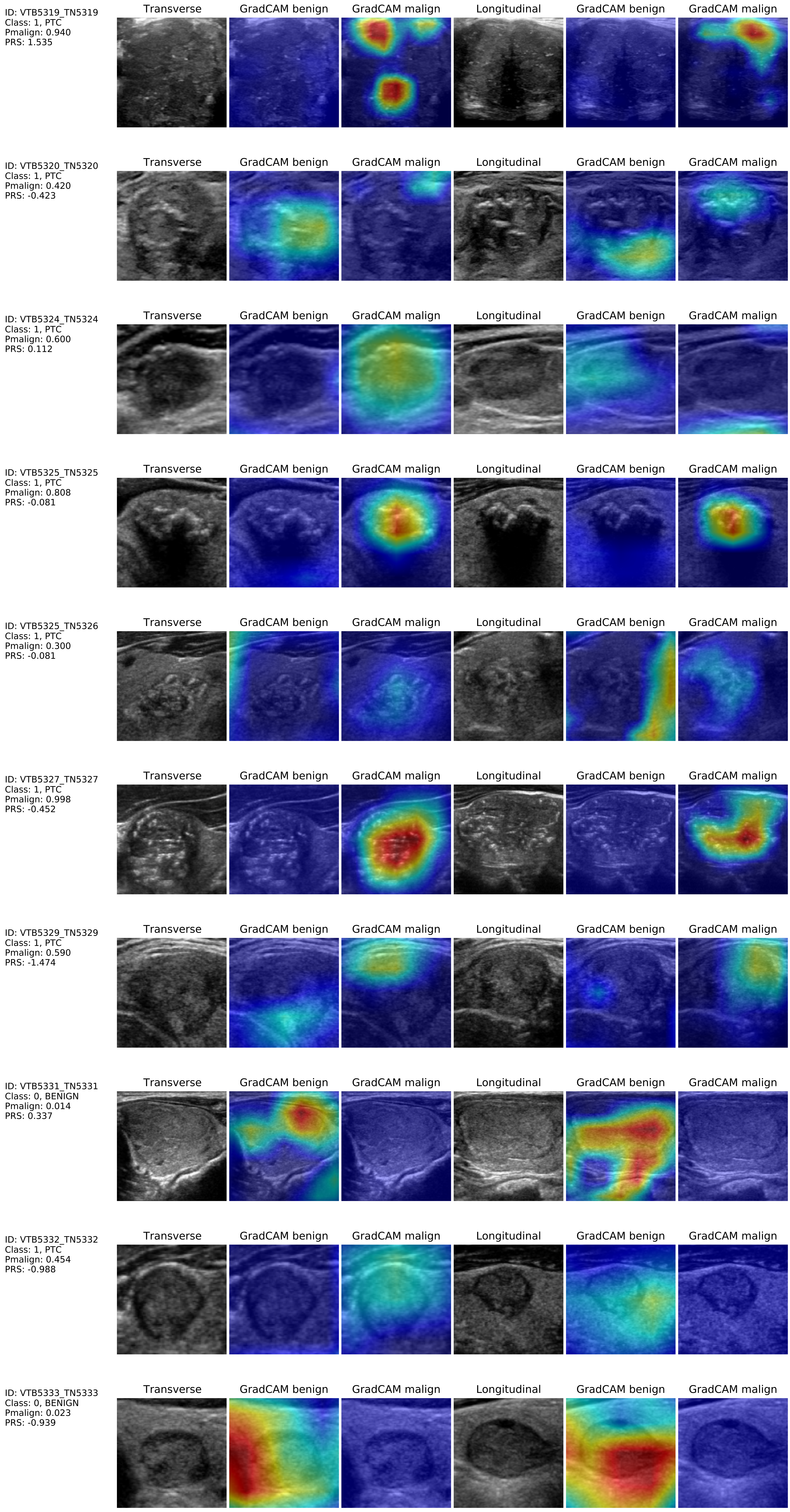

ID: VTB5319_TN5319
Class: 1, PTC
Pmalign: 0.940
PRS: 1.535
Transverse
GradCAM benign
GradCAM malign
Longitudinal
GradCAM benign
GradCAM malign
ID: VTB5320_TN5320
Class: 1, PTC
Pmalign: 0.420
PRS: -0.423
Transverse
GradCAM benign
GradCAM malign
Longitudinal
GradCAM benign
GradCAM malign
ID: VTB5324_TN5324
Class: 1, PTC
Pmalign: 0.600
PRS: 0.112
Transverse
GradCAM benign
GradCAM malign
Longitudinal
GradCAM benign
GradCAM malign
ID: VTB5325_TN5325
Class: 1, PTC
Pmalign: 0.808
PRS: -0.081
Transverse
GradCAM benign
GradCAM malign
Longitudinal
GradCAM benign
GradCAM malign
ID: VTB5325_TN5326
Class: 1, PTC
Pmalign: 0.300
PRS: -0.081
Transverse
GradCAM benign
GradCAM malign
Longitudinal
GradCAM benign
GradCAM malign
ID: VTB5327_TN5327
Class: 1, PTC
Pmalign: 0.998
PRS: -0.452
Transverse
GradCAM benign
GradCAM malign
Longitudinal
GradCAM benign
GradCAM malign
ID: VTB5329_TN5329
Class: 1, PTC
Pmalign: 0.590
PRS: -1.474
Transverse
GradCAM benign
GradCAM malign
Longitudinal
GradCAM benign
GradCAM malign
ID: VTB5331_TN5331
Class: 0, BENIGN
Pmalign: 0.014
PRS: 0.337
Transverse
GradCAM benign
GradCAM malign
Longitudinal
GradCAM benign
GradCAM malign
ID: VTB5332_TN5332
Class: 1, PTC
Pmalign: 0.454
PRS: -0.988
Transverse
GradCAM benign
GradCAM malign
Longitudinal
GradCAM benign
GradCAM malign
ID: VTB5333_TN5333
Class: 0, BENIGN
Pmalign: 0.023
PRS: -0.939
Transverse
GradCAM benign
GradCAM malign
Longitudinal
GradCAM benign
GradCAM malign

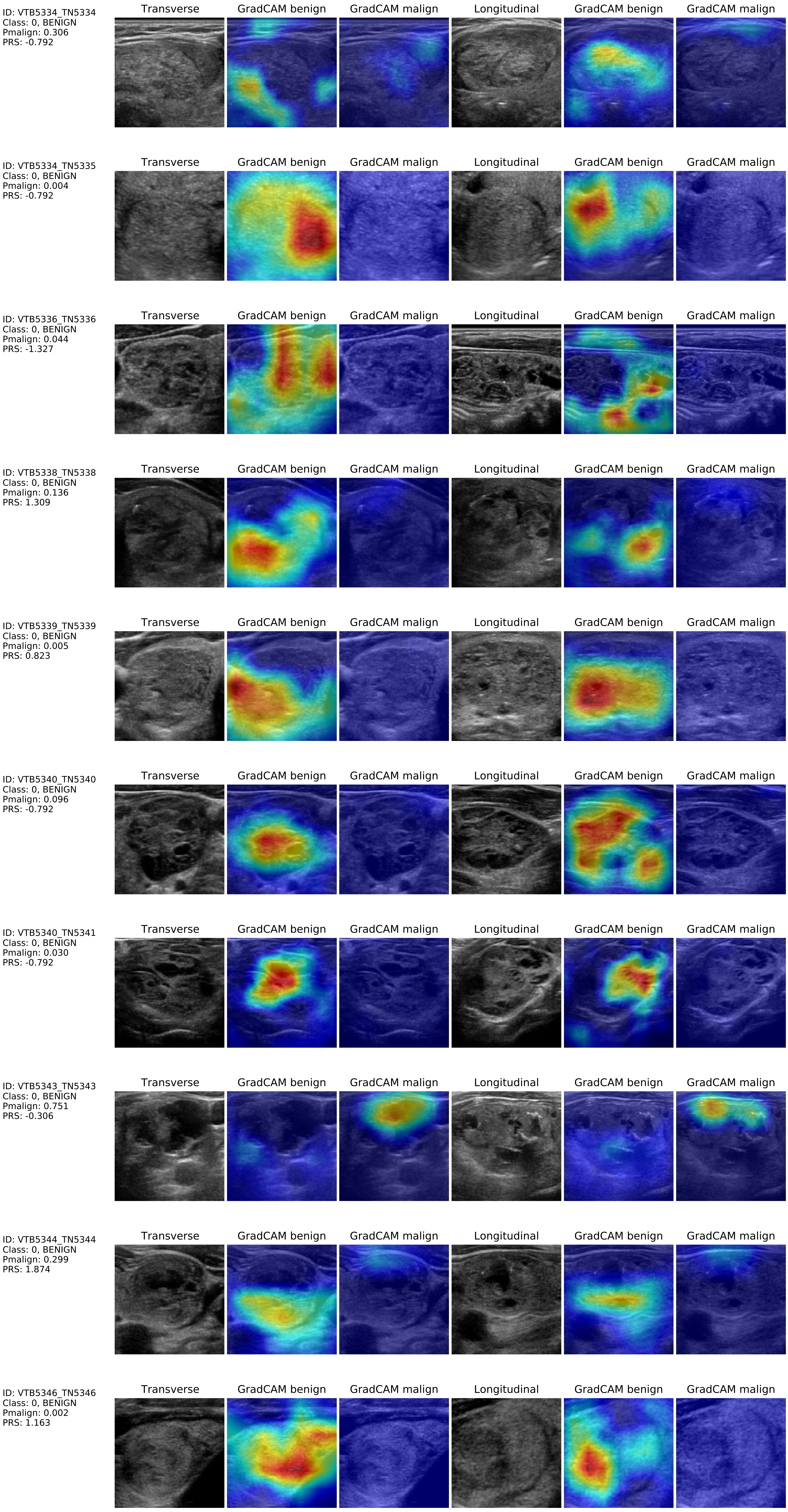
ID: VTB5334_TN5334
Class: 0, BENIGN
Pmalign: 0.306
PRS: -0.792
Transverse
GradCAM benign
GradCAM malign
Longitudinal
GradCAM benign
GradCAM malign
ID: VTB5334_TN5335
Class: 0, BENIGN
Pmalign: 0.004
PRS: -0.792
Transverse
GradCAM benign
GradCAM malign
Longitudinal
GradCAM benign
GradCAM malign
ID: VTB5336_TN5336
Class: 0, BENIGN
Pmalign: 0.044
PRS: -1.327
Transverse
GradCAM benign
GradCAM malign
Longitudinal
GradCAM benign
GradCAM malign
ID: VTB5338_TN5338
Class: 0, BENIGN
Pmalign: 0.136
PRS: 1.309
Transverse
GradCAM benign
GradCAM malign
Longitudinal
GradCAM benign
GradCAM malign
ID: VTB5339_TN5339
Class: 0, BENIGN
Pmalign: 0.005
PRS: 0.823
Transverse
GradCAM benign
GradCAM malign
Longitudinal
GradCAM benign
GradCAM malign
ID: VTB5340_TN5340
Class: 0, BENIGN
Pmalign: 0.096
PRS: -0.792
Transverse
GradCAM benign
GradCAM malign
Longitudinal
GradCAM benign
GradCAM malign
ID: VTB5340_TN5341
Class: 0, BENIGN
Pmalign: 0.030
PRS: -0.792
Transverse
GradCAM benign
GradCAM malign
Longitudinal
GradCAM benign
GradCAM malign
ID: VTB5343_TN5343
Class: 0, BENIGN
Pmalign: 0.751
PRS: -0.306
Transverse
GradCAM benign
GradCAM malign
Longitudinal
GradCAM benign
GradCAM malign
ID: VTB5344_TN5344
Class: 0, BENIGN
Pmalign: 0.299
PRS: 1.874
Transverse
GradCAM benign
GradCAM malign
Longitudinal
GradCAM benign
GradCAM malign
ID: VTB5346_TN5346
Class: 0, BENIGN
Pmalign: 0.002
PRS: 1.163
Transverse
GradCAM benign
GradCAM malign
Longitudinal
GradCAM benign
GradCAM malign

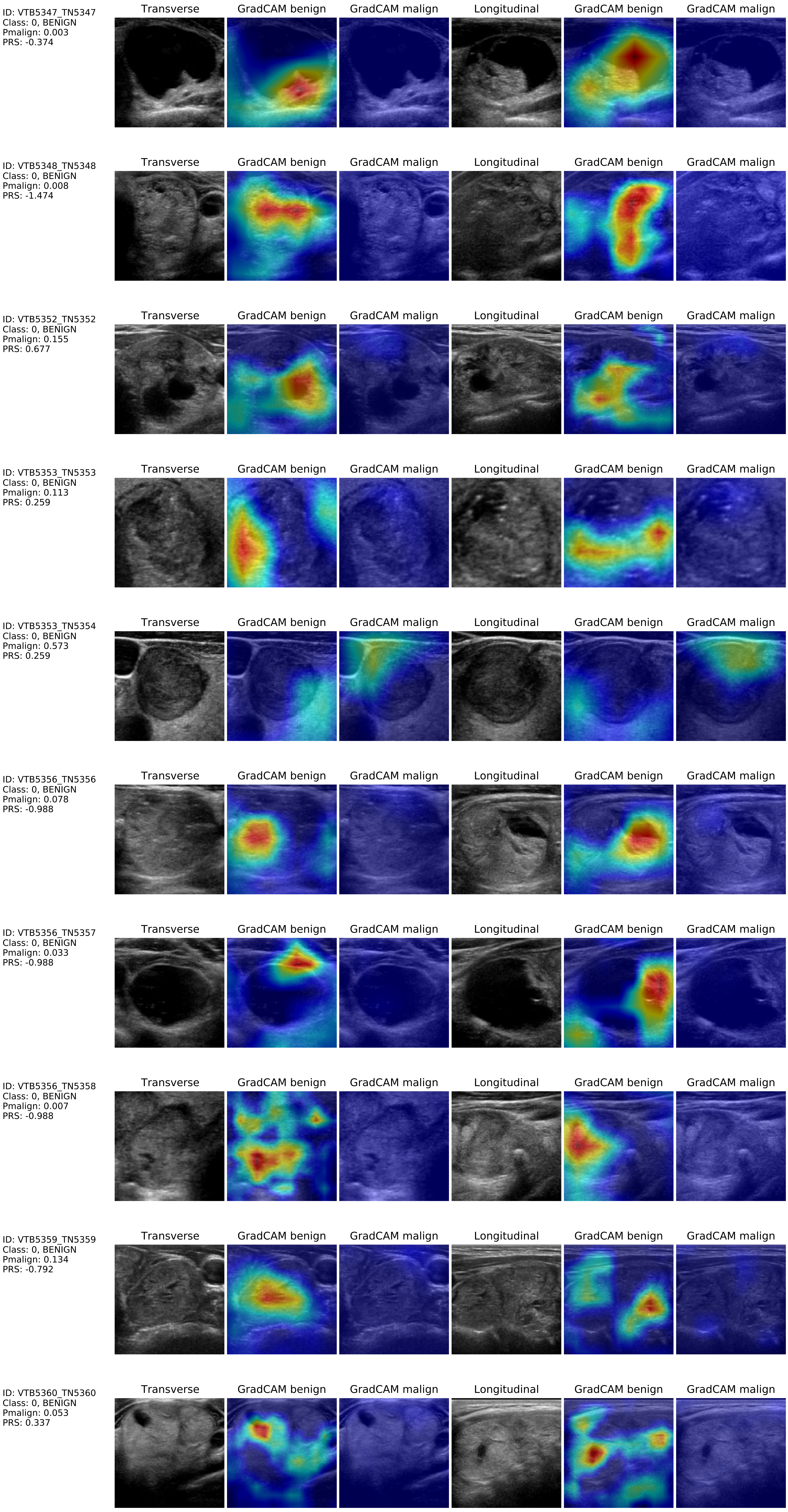
ID: VTB5347_TN5347
Class: 0, BENIGN
Pmalign: 0.003
PRS: -0.374
Transverse
GradCAM benign
GradCAM malign
Longitudinal
GradCAM benign
GradCAM malign
ID: VTB5348_TN5348
Class: 0, BENIGN
Pmalign: 0.008
PRS: -1.474
ID: VTB5352_TN5352
Class: 0, BENIGN
Pmalign: 0.155
PRS: 0.677
ID: VTB5353_TN5353
Class: 0, BENIGN
Pmalign: 0.113
PRS: 0.259
ID: VTB5353_TN5354
Class: 0, BENIGN
Pmalign: 0.573
PRS: 0.259
ID: VTB5356_TN5356
Class: 0, BENIGN
Pmalign: 0.078
PRS: -0.988
ID: VTB5356_TN5357
Class: 0, BENIGN
Pmalign: 0.033
PRS: -0.988
ID: VTB5356_TN5358
Class: 0, BENIGN
Pmalign: 0.007
PRS: -0.988
ID: VTB5359_TN5359
Class: 0, BENIGN
Pmalign: 0.134
PRS: -0.792
ID: VTB5360_TN5360
Class: 0, BENIGN
Pmalign: 0.053
PRS: 0.337

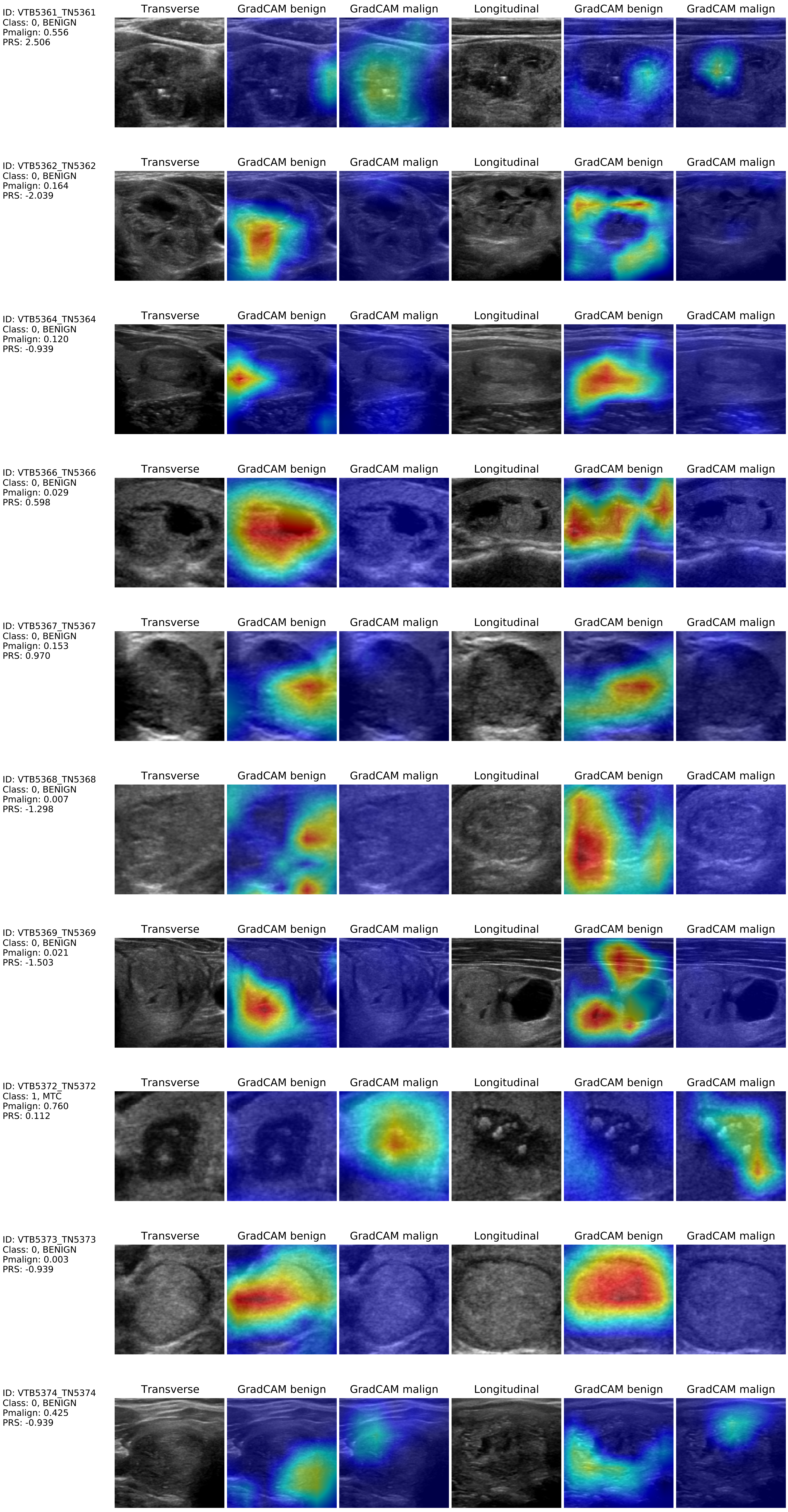
ID: VTB5361_TN5361
Class: 0, BENIGN
Pmalign: 0.556
PRS: 2.506
Transverse
GradCAM benign
GradCAM malign
Longitudinal
GradCAM benign
GradCAM malign
ID: VTB5362_TN5362
Class: 0, BENIGN
Pmalign: 0.164
PRS: -2.039
Transverse
GradCAM benign
GradCAM malign
Longitudinal
GradCAM benign
GradCAM malign
ID: VTB5364_TN5364
Class: 0, BENIGN
Pmalign: 0.120
PRS: -0.939
Transverse
GradCAM benign
GradCAM malign
Longitudinal
GradCAM benign
GradCAM malign
ID: VTB5366_TN5366
Class: 0, BENIGN
Pmalign: 0.029
PRS: 0.598
Transverse
GradCAM benign
GradCAM malign
Longitudinal
GradCAM benign
GradCAM malign
ID: VTB5367_TN5367
Class: 0, BENIGN
Pmalign: 0.153
PRS: 0.970
Transverse
GradCAM benign
GradCAM malign
Longitudinal
GradCAM benign
GradCAM malign
ID: VTB5368_TN5368
Class: 0, BENIGN
Pmalign: 0.007
PRS: -1.298
Transverse
GradCAM benign
GradCAM malign
Longitudinal
GradCAM benign
GradCAM malign
ID: VTB5369_TN5369
Class: 0, BENIGN
Pmalign: 0.021
PRS: -1.503
Transverse
GradCAM benign
GradCAM malign
Longitudinal
GradCAM benign
GradCAM malign
ID: VTB5372_TN5372
Class: 1, MTC
Pmalign: 0.760
PRS: 0.112
Transverse
GradCAM benign
GradCAM malign
Longitudinal
GradCAM benign
GradCAM malign
ID: VTB5373_TN5373
Class: 0, BENIGN
Pmalign: 0.003
PRS: -0.939
Transverse
GradCAM benign
GradCAM malign
Longitudinal
GradCAM benign
GradCAM malign
ID: VTB5374_TN5374
Class: 0, BENIGN
Pmalign: 0.425
PRS: -0.939
Transverse
GradCAM benign
GradCAM malign
Longitudinal
GradCAM benign
GradCAM malign

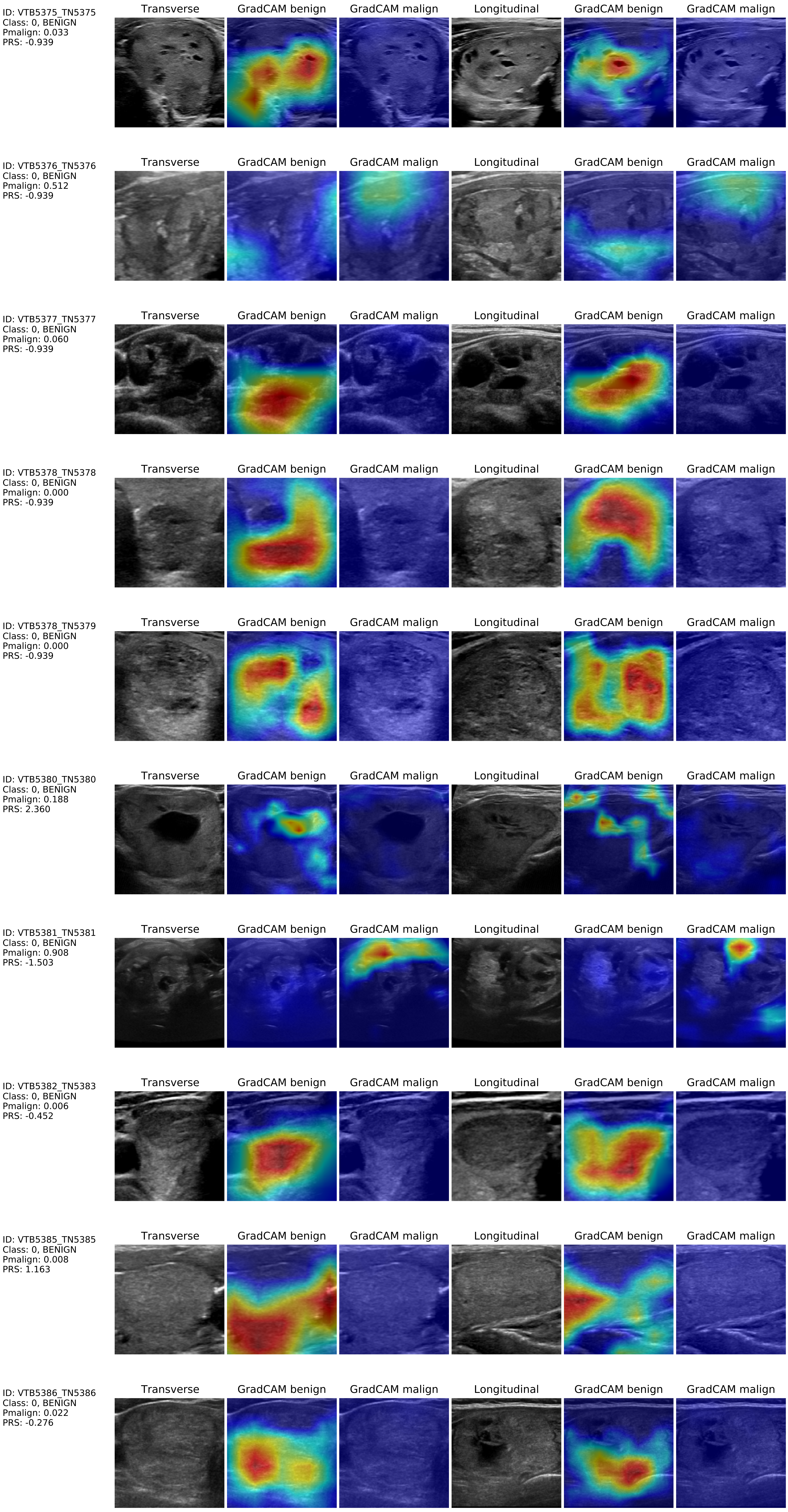
ID: VTB5375_TN5375
Class: 0, BENIGN
Pmalign: 0.033
PRS: -0.939
Transverse
GradCAM benign
GradCAM malign
Longitudinal
GradCAM benign
GradCAM malign
ID: VTB5376_TN5376
Class: 0, BENIGN
Pmalign: 0.512
PRS: -0.939
Transverse
GradCAM benign
GradCAM malign
Longitudinal
GradCAM benign
GradCAM malign
ID: VTB5377_TN5377
Class: 0, BENIGN
Pmalign: 0.060
PRS: -0.939
Transverse
GradCAM benign
GradCAM malign
Longitudinal
GradCAM benign
GradCAM malign
ID: VTB5378_TN5378
Class: 0, BENIGN
Pmalign: 0.000
PRS: -0.939
Transverse
GradCAM benign
GradCAM malign
Longitudinal
GradCAM benign
GradCAM malign
ID: VTB5378_TN5379
Class: 0, BENIGN
Pmalign: 0.000
PRS: -0.939
Transverse
GradCAM benign
GradCAM malign
Longitudinal
GradCAM benign
GradCAM malign
ID: VTB5380_TN5380
Class: 0, BENIGN
Pmalign: 0.188
PRS: 2.360
Transverse
GradCAM benign
GradCAM malign
Longitudinal
GradCAM benign
GradCAM malign
ID: VTB5381_TN5381
Class: 0, BENIGN
Pmalign: 0.908
PRS: -1.503
Transverse
GradCAM benign
GradCAM malign
Longitudinal
GradCAM benign
GradCAM malign
ID: VTB5382_TN5383
Class: 0, BENIGN
Pmalign: 0.006
PRS: -0.452
Transverse
GradCAM benign
GradCAM malign
Longitudinal
GradCAM benign
GradCAM malign
ID: VTB5385_TN5385
Class: 0, BENIGN
Pmalign: 0.008
PRS: 1.163
Transverse
GradCAM benign
GradCAM malign
Longitudinal
GradCAM benign
GradCAM malign
ID: VTB5386_TN5386
Class: 0, BENIGN
Pmalign: 0.022
PRS: -0.276
Transverse
GradCAM benign
GradCAM malign
Longitudinal
GradCAM benign
GradCAM malign

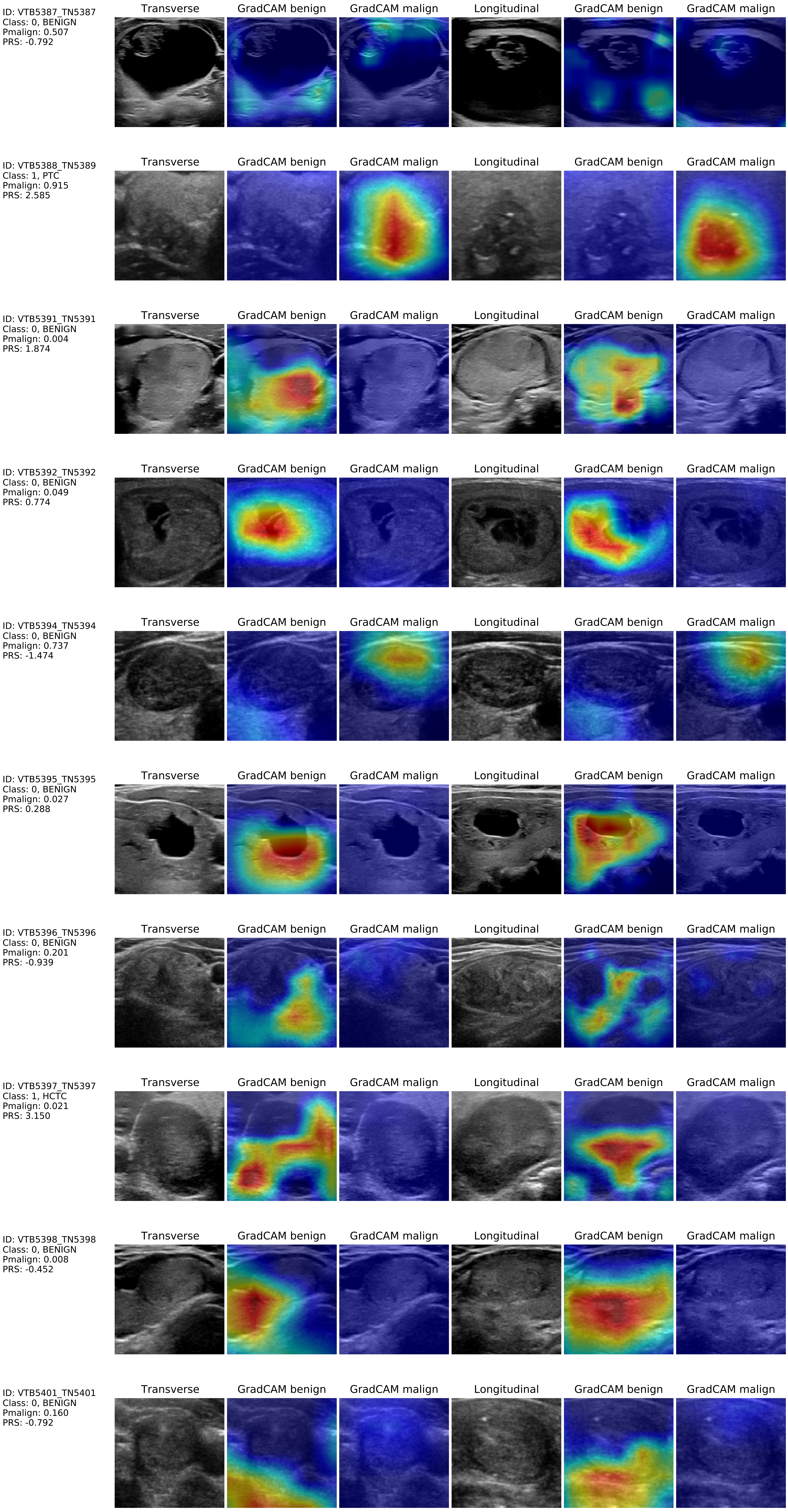
ID: VTB5387_TN5387
Class: 0, BENIGN
Pmalign: 0.507
PRS: -0.792
Transverse
GradCAM benign
GradCAM malign
Longitudinal
GradCAM benign
GradCAM malign
ID: VTB5388_TN5389
Class: 1, PTC
Pmalign: 0.915
PRS: 2.585
Transverse
GradCAM benign
GradCAM malign
Longitudinal
GradCAM benign
GradCAM malign
ID: VTB5391_TN5391
Class: 0, BENIGN
Pmalign: 0.004
PRS: 1.874
Transverse
GradCAM benign
GradCAM malign
Longitudinal
GradCAM benign
GradCAM malign
ID: VTB5392_TN5392
Class: 0, BENIGN
Pmalign: 0.049
PRS: 0.774
Transverse
GradCAM benign
GradCAM malign
Longitudinal
GradCAM benign
GradCAM malign
ID: VTB5394_TN5394
Class: 0, BENIGN
Pmalign: 0.737
PRS: -1.474
Transverse
GradCAM benign
GradCAM malign
Longitudinal
GradCAM benign
GradCAM malign
ID: VTB5395_TN5395
Class: 0, BENIGN
Pmalign: 0.027
PRS: 0.288
Transverse
GradCAM benign
GradCAM malign
Longitudinal
GradCAM benign
GradCAM malign
ID: VTB5396_TN5396
Class: 0, BENIGN
Pmalign: 0.201
PRS: -0.939
Transverse
GradCAM benign
GradCAM malign
Longitudinal
GradCAM benign
GradCAM malign
ID: VTB5397_TN5397
Class: 1, HCTC
Pmalign: 0.021
PRS: 3.150
Transverse
GradCAM benign
GradCAM malign
Longitudinal
GradCAM benign
GradCAM malign
ID: VTB5398_TN5398
Class: 0, BENIGN
Pmalign: 0.008
PRS: -0.452
Transverse
GradCAM benign
GradCAM malign
Longitudinal
GradCAM benign
GradCAM malign
ID: VTB5401_TN5401
Class: 0, BENIGN
Pmalign: 0.160
PRS: -0.792
Transverse
GradCAM benign
GradCAM malign
Longitudinal
GradCAM benign
GradCAM malign

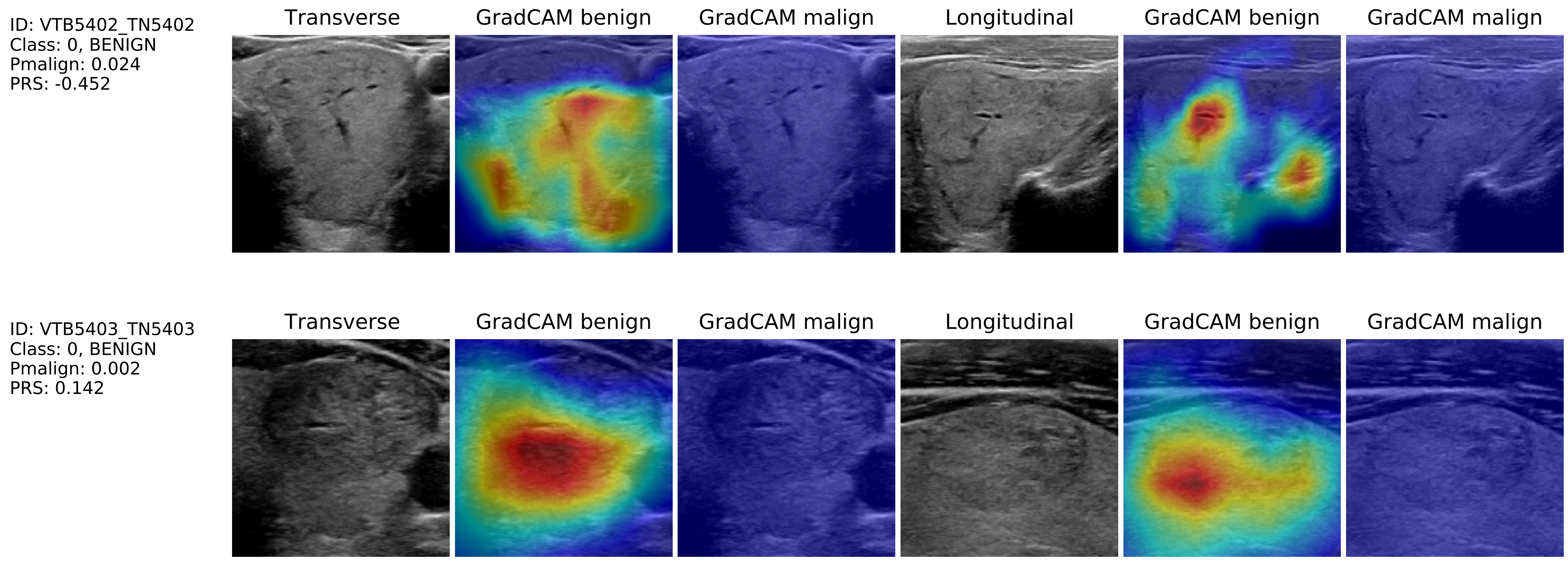
ID: VTB5402_TN5402
Class: 0, BENIGN
Pmalign: 0.024
PRS: -0.452
Transverse
GradCAM benign
GradCAM malign
Longitudinal
GradCAM benign
GradCAM malign
ID: VTB5403_TN5403
Class: 0, BENIGN
Pmalign: 0.002
PRS: 0.142
Transverse
GradCAM benign
GradCAM malign
Longitudinal
GradCAM benign
GradCAM malign
